# A brain-persistent DDR2-degrading antibody reverses Alzheimer’s pathologies by restoring brain fluid dynamics and metabolic clearance

**DOI:** 10.64898/2026.03.17.26348575

**Authors:** Penghui Yang, Xu Chen, Jiuyang Ding, Yuzhou Peng, Zhixiang Lei, Ling Mou, Yunpeng Xiao, Yunxin Lai, Tianyang Guo, Junqi Li, Jinjun Jiang, Xihui Huang, Guilin Li, Xiaoting Lin, Su Feng, Bing Su, Chuhui Gao, Jiguo Liu, Feng Qian, Haishan Jiang, Pengcheng Ran, Yaxin Li, Jia Li, Jin Su

## Abstract

Alzheimer’s disease (AD) is defined by Aβ deposition, yet cerebrovascular and glymphatic dysfunction are early drivers of progression. We identify discoidin domain receptor 2 (DDR2), a collagen-sensing receptor tyrosine kinase, as a central mediator of neurovascular impairment in AD. Integrative analysis of human single-nucleus RNA (snRNA)-seq data and immunohistochemical validation across human, non-human primate, and mouse AD models revealed that DDR2 is markedly upregulated in reactive astrocytes, perivascular fibroblasts (PVFs), and choroid plexus epithelial cells (CPECs). Astrocytic DDR2 overexpression in APP/PS1 mice exacerbated Aβ deposition, reduces cerebral blood flow (CBF), and impaired glymphatic function. In CPECs, DDR2 upregulation accompanied increased type X collagen (Collagen X). Central nervous system (CNS) delivery of a monoclonal antibody (HL2) that promotes lysosomal degradation of DDR2 ablated pathogenic signaling, reduced Collagen X, and rescued cognitive, vascular, and glymphatic deficits in APP/PS1 mice. A DDR2-targeted PET probe enabled *in vivo* visualization of target engagement in these models. These findings establish DDR2 as a key driver of neurovascular dysfunction and support CNS-delivered DDR2-targeted therapy for AD.

**INTRODUCTION:** Alzheimer’s disease (AD) is defined by amyloid-β (Aβ) plaques and tau tangles, yet the modest clinical benefits of recently approved immunotherapies underscore its multifactorial nature. Cerebrovascular dysfunction and impaired glymphatic clearance are now recognized as early and integral contributors to AD pathogenesis, but the molecular drivers disrupting these vascular and clearance networks remain poorly defined.

**RATIONALE:** Discoidin domain receptor 2 (DDR2), a collagen-binding receptor tyrosine kinase, drives fibrosis in peripheral organs and has been linked to AD pathology, yet the specific cell types and mechanisms through which DDR2 contributes to disease progression remain unknown. Furthermore, strategies for sustained *in vivo* suppression of DDR2, together with tools for non-invasive monitoring, are lacking. We therefore mapped DDR2 expression across human AD brains, modeled its overexpression in mice, and developed a targeted therapeutic and imaging strategy.

**RESULTS:** Integrative analysis of human single-nucleus RNA-seq data and immunohistochemical validation across human, non-human primate, and mouse AD brains revealed pronounced DDR2 upregulation in three functionally distinct cell populations: reactive astrocytes, perivascular fibroblasts (PVFs), and choroid plexus epithelial cells (CPECs). In astrocytes, DDR2 expression correlated positively with Braak stage, Aβ burden, and cognitive decline. Astrocyte specific *Ddr2* overexpression in APP/PS1 mice exacerbated Aβ deposition via β-amyloid cleaving enzyme 1 (BACE1) upregulation, reduced cerebral blood flow (CBF), disrupted blood-brain barrier (BBB) integrity, and impaired glymphatic function. In CPECs, DDR2 upregulation was accompanied by increased expression of type X collagen (Collagen X), a non-fibrillar collagen that is both a DDR2 activator and a marker of calcification, suggesting a potential link to choroid plexus calcification. We further developed a monoclonal antibody (HL2) that drives efficient internalization and lysosomal degradation of DDR2 regardless of collagen occupancy, by binding a unique “waist” epitope distinct from the collagen-binding site. Adeno-associated virus (AAV)-mediated central nervous system (CNS) delivery of HL2 in APP/PS1 mice ablated DDR2, reduced Collagen X expression, reversed cognitive impairment, diminished Aβ plaque burden. This was accompanied by restored cerebral perfusion, enhanced glucose supply, and normalized CSF dynamics. A DDR2-targeted PET tracer, validated in human idiopathic pulmonary fibrosis (IPF) studies, enabled *in vivo* visualization of target engagement in mouse AD models.

**CONCLUSION:** This study identifies DDR2 as a master regulator of multi-compartmental failure in AD, orchestrating dysfunction across cerebral vasculature, CSF dynamics, and the choroid plexus. We developed a monoclonal antibody that drives durable internalization and lysosomal degradation of DDR2; when delivered via a neuron-tropic AAV, it achieves sustained CNS expression and confers broad therapeutic efficacy against AD. A companion DDR2-targeted PET tracer enables noninvasive visualization of target engagement in vivo. Together, this integrated platform establishes a new paradigm for disease-modifying therapy in AD.

DDR2 drives multi-compartmental dysfunction in Alzheimer’s disease.
The upper region depicts an AD brain where DDR2 is upregulated in astrocytes, PVFs, and CPECs. DDR2-activating ligands Collagen I and Collagen X are highly expressed in PVFs and CPECs, respectively. In astrocytes and PVFs, DDR2 impairs CBF and energy supply, disrupts CSF circulation, and promotes Aβ deposition. In CPECs, DDR2 induces Collagen X expression, linking to choroid plexus dysfunction. The middle region shows therapeutic intervention with AAV-delivered HL2, a monoclonal antibody that binds a unique epitope on DDR2 non-competitive with collagen binding and promotes its lysosomal degradation, thereby rescuing cognitive and vascular deficits. The site of action of DDR2 blockade by HL2 is also depicted.

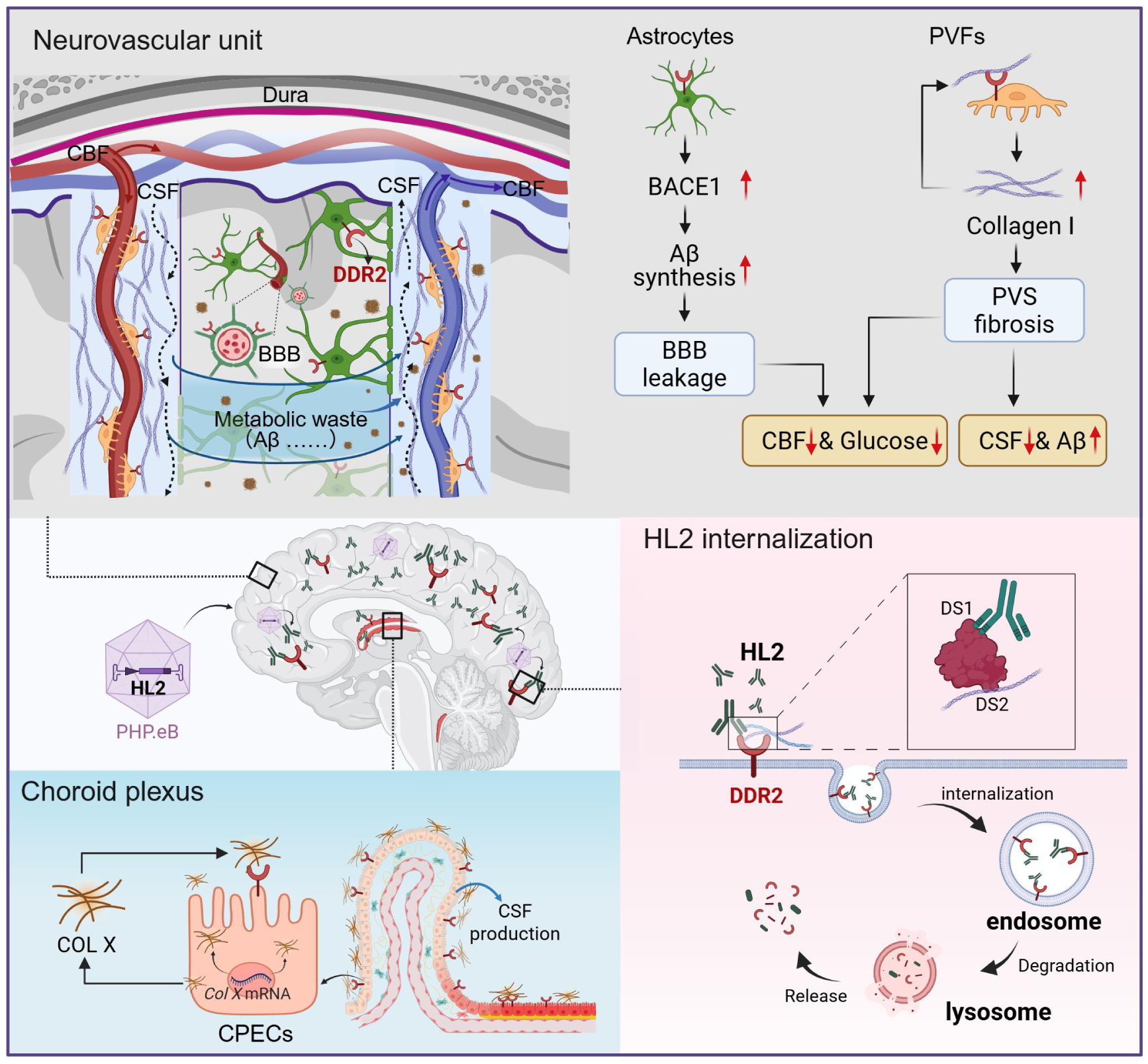

## Introduction

Alzheimer’s disease (AD) is defined by amyloid-β (Aβ) plaques and tau tangles, yet the modest clinical benefits of recently approved immunotherapies underscore its multifactorial nature and the urgent need for complementary strategies (*1*). Attention has increasingly turned to non-neuronal mechanisms, particularly cerebrovascular dysfunction and impaired glymphatic clearance, now recognized as early and integral contributors to AD pathogenesis (*2–7*). However, the molecular drivers disrupting these vascular and clearance networks remain poorly defined, representing a critical knowledge gap.

Discoidin domain receptor 2 (DDR2), a collagen-binding receptor tyrosine kinase, is well known for its pro-fibrotic role in peripheral diseases (*8–12*). Aberrant perivascular fibrosis has recently emerged as a feature of AD (*13*), and acute DDR2 knockdown has been shown to mitigate AD-like pathology (*14*). Yet, the specific cell types and mechanisms through which DDR2 contributes to AD progression remain unknown. Moreover, strategies for sustained *in vivo* suppression of DDR2, together with tools for non-invasive monitoring, are lacking.

Here, we identify DDR2 upregulation in astrocytes, perivascular fibroblasts (PVFs), and choroid plexus epithelial cells (CPECs) as a hallmark of AD pathology, with expression levels correlating to amyloid burden and cognitive decline. Using astrocyte specific overexpression in mice, we establish a causal role for astrocytic DDR2 in driving neurovascular dysfunction and AD-like pathology. We further demonstrate that sustained central nervous system (CNS) delivery of a monoclonal antibody (HL2), which binds a non-collagen-interfering epitope and promotes lysosomal degradation of DDR2, rescues cognitive function and restores cerebral and glymphatic perfusion.

## Results

### DDR2 is upregulated across astrocytes, PVFs, and choroid plexus epithelium in AD

To identify the cell types expressing *DDR2* in AD brains, we first analyzed public single-cell RNA-sequencing (scRNA-seq) data from the AD Multi-Region Database (*15*). We found that *DDR2* is mainly expressed in astrocytes, fibroblasts, and CPECs (Fig. 1A). Re-clustering of astrocytes revealed pronounced enrichment of *DDR2^+^* astrocytes in samples with higher Braak staging (Fig. 1B, and fig. S1, A to D). In AD astrocytes, *DDR2* expression positively correlated with 34 established astrocyte biomarkers (fig. S1, E and F) (*16*). We developed HL2, a monoclonal antibody that specifically recognizes the native DDR2 extracellular domain without cross-reacting with human or mouse DDR1 (fig. S2, A to E). Its performance in tissue immunofluorescence was confirmed on brain sections from AAV-mDDR2-overexpressing mice, where it robustly labeled DDR2 in situ, demonstrating its utility for detecting endogenous brain protein (fig. S2, F to H). Immunostaining of human cortical sections using HL2 and the astrocyte marker glial fibrillary acidic protein (GFAP) revealed upregulated DDR2 expression in AD astrocytes (Fig. 1C). Similarly, APP/PS1 mice showed elevated DDR2 co-localized with GFAP in parenchymal and perivascular astrocytes (fig. S3A), alongside Aβ accumulation and neuronal loss (fig. S3, B and C). In Aβ oligomer (Aβ)-treated cynomolgus monkeys (*17*), DDR2 also co-localized with GFAP in the frontal cortex (fig. S3D), and its expression correlated with Aβ pathology across models (fig. S3E). Together, these findings demonstrate conserved upregulation of astrocytic DDR2 across species in AD, closely associated with Aβ pathology.

**Fig. 1.**
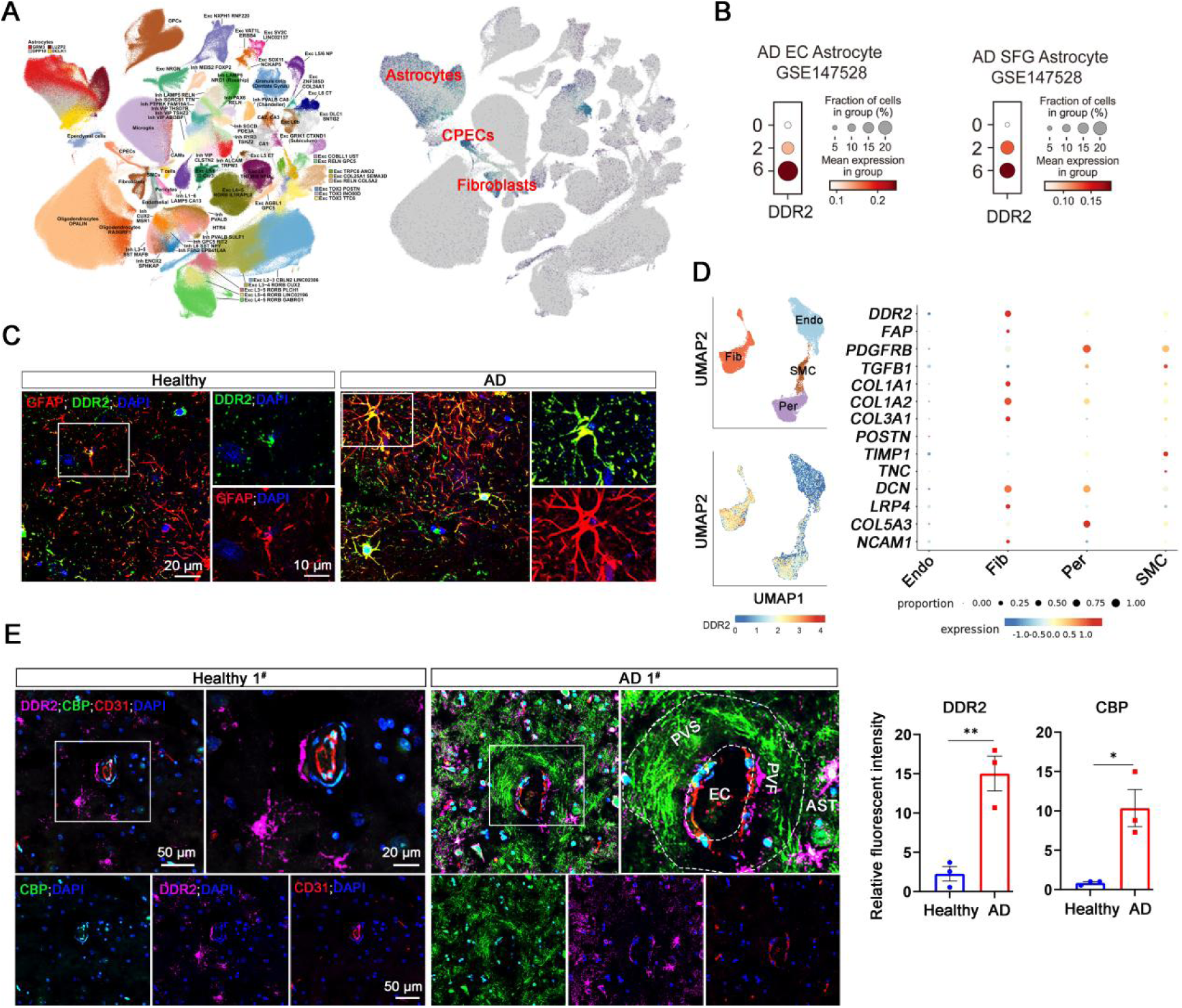
DDR2 expression in astrocytes and PVFs in human AD brains. **(A)** UMAP plots showing 75 identified cell types in human AD brain (*left*), and the pattern of DDR2 expression in these cell types, highlighting the 3 cell types with the highest DDR2 expression (*right*). **(B)** Dot plots showing DDR2 expression levels in astrocytes from EC or SFG of AD patients at different Braak stages. **(C)** Immunofluorescence staining for DDR2 (green) and GFAP (red) was performed on frozen brain sections from a healthy donor and an AD patient. The areas outlined by white squares are magnified in the adjacent panels, with separate channels shown for GFAP and DDR2 signals (Scale bar: 20 μm for original images and 10 μm for enlarged images). **(D)** UMAP plots showing DDR2 expression pattern specifically among endothelial cells, fibroblasts, pericytes and smooth muscle cells from human AD brains (*left*), with dot plots showing the expression levels of fibrosis-related marker genes in these four cell types (*right*). **(E)** Representative immunofluorescence images showing DDR2 (purple), CD31 (red) and collagen (green) in the cerebral cortex from a healthy donor and an AD patient, the areas outlined by white squares are magnified in the adjacent panels (*left*: Scale bar: 50 μm for original images and 20 μm for enlarged images). Quantification of brain DDR2 and CBP signal intensity (*right*). White dashed lines outline blood vessels and PVSs, respectively.

Fibroblasts, marked by expression of the pan-fibroblastic gene decorin (*DCN)* and fibrillar collagens (*COL1A1, COL1A2, COL3A1*), represent a major *DDR2* expressing population in the brain vasculature (Fig. 1D). To visualize fibrotic foci, we applied a collagen-binding peptide (CBP) previously validated for in vivo imaging of Collagen I in pulmonary fibrosis (*18, 19*). The binding affinity of FITC-conjugated CBP against Collagen I was validated by siRNA-mediated knockdown (fig. S4, A and B). In healthy aged brain tissue, Collagen I and DDR2 were largely undetectable. In contrast, AD patient brains exhibited marked Collagen I deposition surrounding DDR2 positive cells in the cortex and hippocampus. These DDR2 positive cells displayed spindle-shaped morphology consistent with PVFs and closely enveloped large vessels (Fig. 1E; fig. S5, A and B). Together with astrocytes, DDR2 positive PVFs delineated perivascular spaces (PVSs) densely filled with Collagen I, suggesting that perivascular fibrosis contributes to neurovascular dysfunction in AD. In AD mouse brains, DCN expression was perivascular and colocalized with DDR2 (fig. S3F), mirroring the perivascular enrichment of DDR2 observed in humans (fig. S3A). Large field sections from human brains confirmed DDR2 positive cells encircling CD31 positive vessels, with pronounced perivascular accumulation in AD cases compared to controls (fig. S5C). Aβ deposition was confirmed in the same tissues (fig. S5D). Human sample characteristics are detailed in table S1, and the research involving human tissue was approved by the Ethics Committee of Guizhou Medical University (approval no. 2023-91).

To characterize collagen expression in CPECs, the CSF-producing epithelium, we analyzed single-cell transcriptomic data using transthyretin (TTR) as a lineage marker. CPECs predominantly expressed COL10A1, a non-fibrillar collagen and the cognate ligand of DDR2 (*20*), with minimal expression of fibrillar collagens COL1A1 or COL3A1 (fig. S6A). Immunostaining confirmed elevated DDR2 and Collagen X in choroid plexus from AD patients and APP/PS1 mice (fig. S6, B-D).

Taken together, these data suggest that DDR2-driven pathology in PVFs, astrocytes, and CPECs may collectively contribute to disruptions in cerebral blood flow (CBF) and CSF homeostasis, positioning multi-compartmental dysfunction as a potential feature of AD progression that warrants further functional investigation.

### Astrocytic DDR2 overexpression exacerbates amyloid pathology and cognitive decline in AD mice

To investigate the functional role of astrocytic DDR2 in AD pathology, we specifically overexpressed mouse *Ddr2* (mDdr2) in astrocytes of 10-month-old APP/PS1 mice using an AAV-PHP.eB vector driven by the *Gfap* promoter, following the experimental timeline illustrated in Fig. 2A. In vivo PET/CT imaging with a DDR2-targeted tracer (developed by our recent study, https://medrxiv.org/cgi/content/short/2025.11.26.25341068v1), revealed elevated DDR2 levels in AD mice at two months post injection, which was further augmented by viral-mediated overexpression (Fig. 2B). At three months, APP/PS1 mice overexpressing astrocytic DDR2 exhibited a marked increase in ¹⁸F-AV45 PET signal, indicating exacerbated amyloid pathology (Fig. 2C). Behavioral assessment in the Morris water maze revealed that these mice had significantly prolonged escape latency and fewer platform crossings compared to wild-type controls, with DDR2 overexpression further impairing performance by ∼45% and ∼65%, respectively (Fig. 2D). Together, these findings demonstrate that astrocytic DDR2 elevation aggravates both amyloid deposition and cognitive decline in a mouse model of AD mice.

**Fig. 2.**
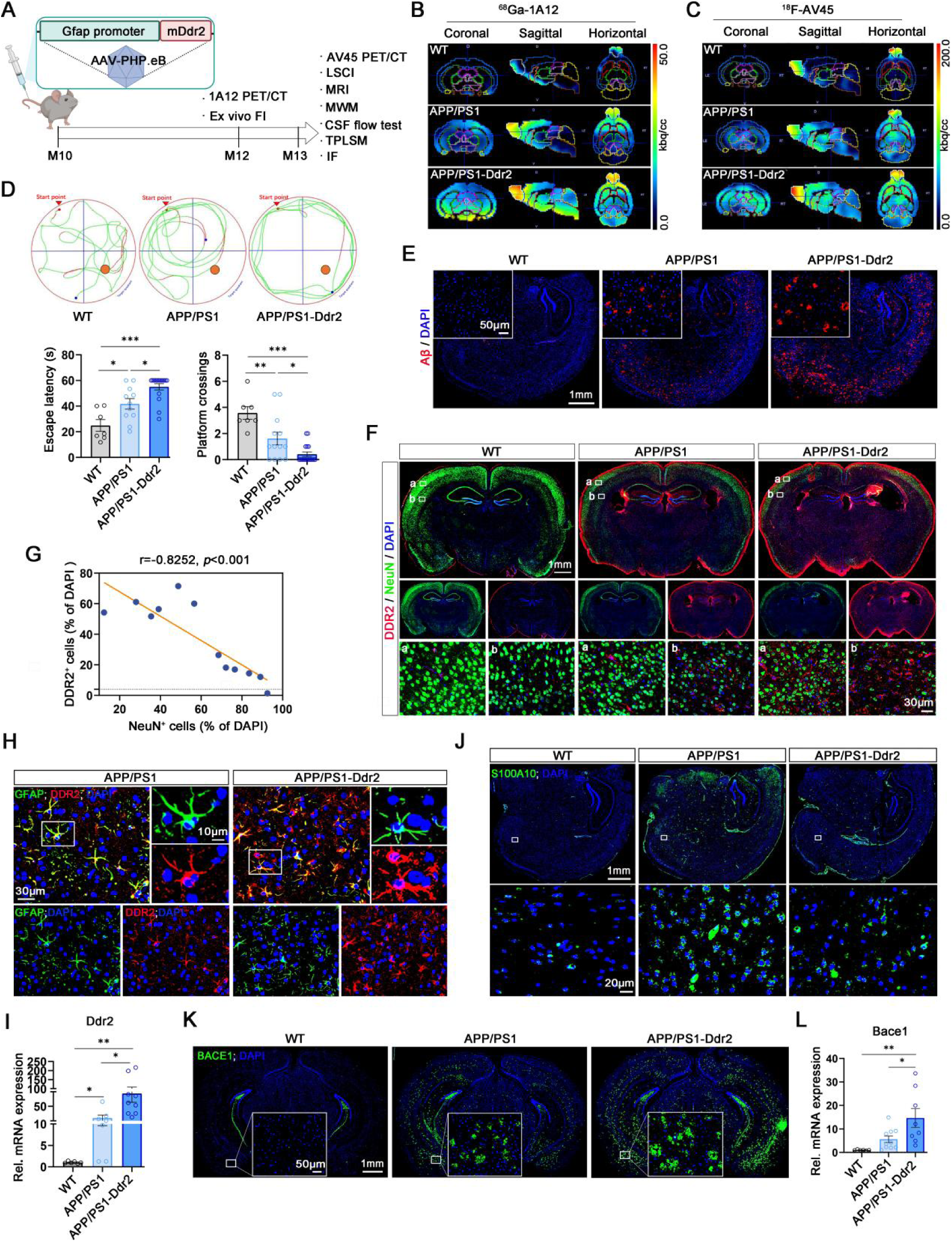
Astrocyte specific overexpression of DDR2 modulates AD pathology in APP/PS1 mice. **(A)** Experimental timeline. Ten month old APP/PS1 mice were injected with an AAV-PHP.eB vector carrying astrocyte specific (*Gfap* promoter) mouse Ddr2. *In vivo* 1A12 PET/CT imaging was performed at 2 months post injection, followed by behavioral tests, AV45 PET/CT imaging, LSCI and tissue collection at 3 months. **(B)** Representative multi planar PET/CT sections depicting regional tracer uptake: coronal (*left*), sagittal (*middle*), and horizontal (*right*) views *in vivo* PET/CT imaging using 1A12. Color scale indicates standardized uptake intensity. **(C)** *In vivo* AV45-PET imaging of amyloid burden. **(D)** Representative motion tracks of APP/PS1 and APP/PS1-mDdr2 transgenic mice in morris water maze performance (*top*); Escape latency during training days (*bottom left*) and the number of platform crossings during the trial (*bottom right*). Data were shown as mean ± SEM and one-way ANOVA analysis was used for data analysis (\**p* <0.05, \*\**p* <0.001, \*\*\**p* <0.0001). **(E)** Representative images of Aβ plaques in the cortex (Scale bar: 1 mm for original images and 50 μm for enlarged images). **(F)** Representative images of DDR2 and NeuN staining in the coronal section of a mouse brain (Scale bar: 1 mm for original images and 30 μm for enlarged images). **(G)** Correlation analysis between DDR2^+^ and NeuN^+^ cells density in cortical regions (Pearson r = −0.8252, *p*<0.001). **(H)** Representative immunofluorescence images of DDR2 (red) and GFAP (green) in cortical brain sections (Scale bar: 30 μm for original images and 10 μm for enlarged images). The areas outlined by white squares are magnified in the adjacent panels. **(I)** qPCR analysis of *Ddr2* mRNA levels in cortical homogenates (Mean ± SEM; \*\**p*<0.01, one-way ANOVA). **(J)** Immunofluorescence staining of the A2 astrocyte marker S100A10 (green) in the cortex (Scale bar: 20 μm). **(K)** Representative immunofluorescence of BACE1 expression in the coronal section of a mouse brain (Scale bar: 1 mm for original images and 50 μm for enlarged images). **(L)** qPCR analysis of *Bace1* mRNA levels in the mouse brain homogenates (Mean ± SEM; \**p*<0.05, \*\**p*<0.01, one-way ANOVA).

Consistent with the above PET and behavioral findings, astrocyte-specific DDR2 overexpression in APP/PS1 mice exacerbated Aβ plaque deposition (Fig. 2E) and neuronal loss (Fig. 2F), with a negative correlation between DDR2 and NeuN signals (Fig. 2G). While GFAP expression remained unchanged (Fig. 2, H and I), DDR2 overexpression selectively downregulated the A2 astrocyte marker S100 calcium binding protein A10 (S100A10) (Fig. 2J), with no effect on the A1 marker complement component 3 (C3) (data not shown). Mechanistically, DDR2 overexpression upregulated BACE1, the rate limiting protease for Aβ generation (*21*), at both the protein and transcriptional levels (Fig. 2, K and L), linking astrocytic DDR2 to enhanced Aβ generation. Interestingly, astrocyte-specific DDR2 overexpression did not alter DDR2 levels in CPECs but markedly increased their Collagen X expression (fig. S6D), indicating that astrocytic DDR2 signaling may remotely influence CPEC biology.

In summary, astrocyte-specific DDR2 overexpression in APP/PS1 mice exacerbated cognitive deficits, promoted Aβ deposition via BACE1 upregulation, and intensified neuronal loss while suppressing protective A2 astrocyte markers. These findings implicate astrocytic DDR2 in driving AD pathogenesis through dual effects on amyloid metabolism and neuroinflammation.

### Astrocytic DDR2 overexpression impairs cerebrovascular function and CSF dynamics

Given the high expression of DDR2 in key constituents of the neurovascular unit, we assessed the CBF in AD mice with astrocyte-specific DDR2 overexpression. Laser speckle contrast imaging (LSCI) revealed that AD mice exhibited reduced CBF compared to wild-type controls, consistent with previous reports (*4, 22*), which was further exacerbated by DDR2 overexpression (Fig. 3A). CBF levels correlated negatively with DDR2 mRNA expression (Fig. 3B).

**Fig. 3.**
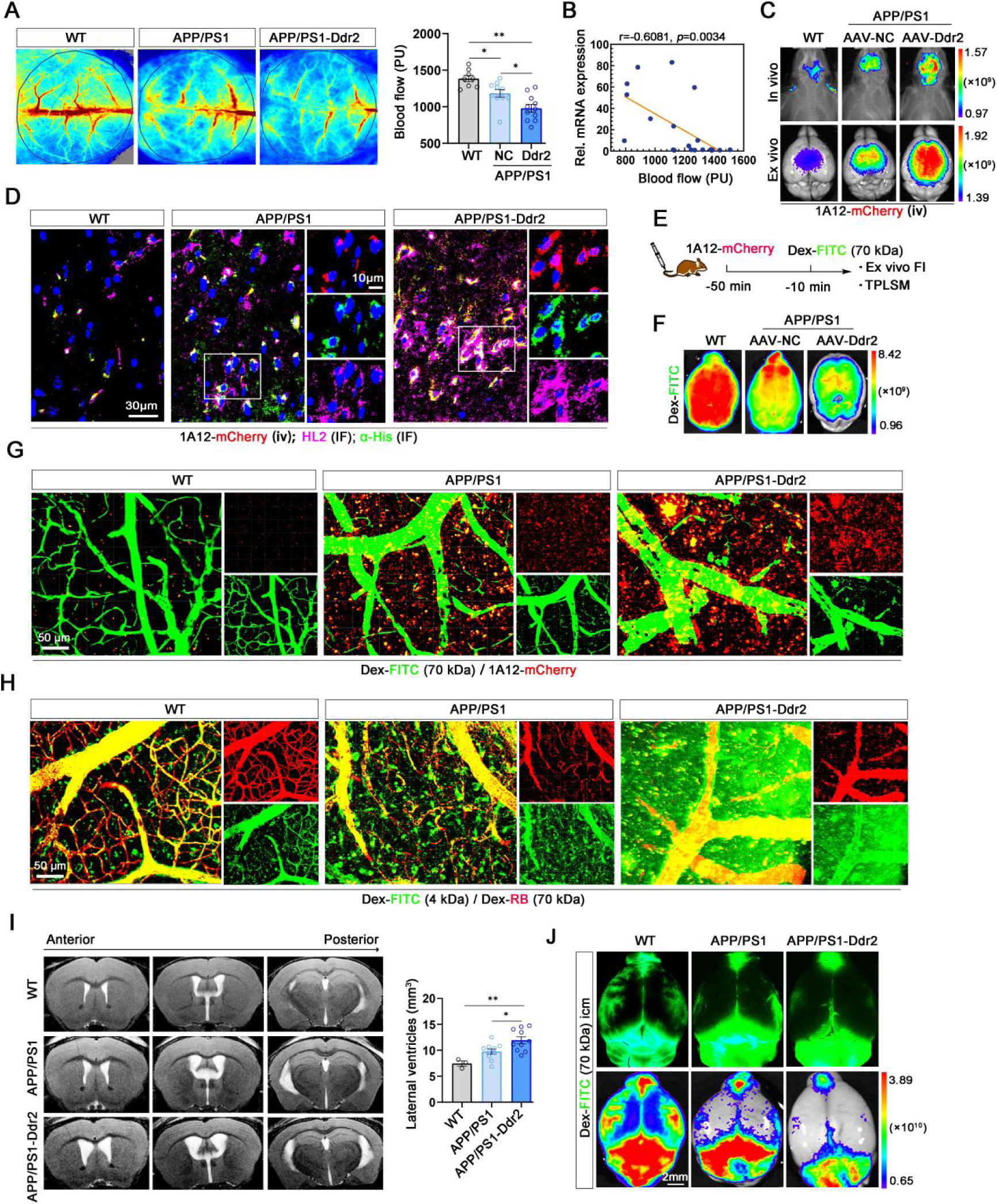
Effect of DDR2 in cerebrovascular dysfunction and CSF dynamics in AD mice. **(A)** CBF assessed by LSCI in WT, APP/PS1, and astrocyte specific Ddr2 overexpressing APP/PS1 mice. Left: Representative pseudocolor perfusion maps. Right: Quantification of relative perfusion units (Mean ± SEM; \*\**p* < 0.01, one-way ANOVA). **(B)** Correlation between CBF (from A) and cortical *Ddr2* mRNA levels (qPCR) across individual mice (Pearson r = −0.6081, *p* < 0.01). **(C)** *In vivo* (top) and *ex vivo* (bottom) whole brain fluorescence imaging after intravenous injection of the DDR2 targeting probe 1A12-mCherry. **(D)** Validation of 1A12-mCherry brain delivery and target specificity: *Ex vivo* brain sections for intrinsic mCherry fluorescence (red), anti-His tag immunofluorescence staining (green), and anti-DDR2 antibody HL2 staining (purple), the areas outlined by white squares are magnified in the adjacent panels. Scale bar: 30 μm for original images and 10 μm for enlarged images. **(E)** Schematic of the sequential probe injection protocol for vascular perfusion assessment: 1A12-mCherry followed 40 min later by Dextran-FITC (70 kDa). **(F)** Whole brain fluorescence imaging of vascular perfusion with Dextran-FITC (70 kDa). **(G)** Two photon microscopy of cortical vasculature. Representative images show mCherry signal (red) and dextran-FITC vasculature (green) in WT, APP/PS1, and APP/PS1-DDR2 mice, scale bars: 50 μm. **(H)** Representative two-photon microscopy images of vascular leakage after injection of dextran-FITC (4 kDa, green) and dextran-RB (70 kDa, red), scale bars: 50 μm. **(I)** Ventricular morphology analyzed by MRI based volumetric reconstruction. Left: Representative images of lateral ventricles from each group. Right: Quantification of lateral ventricular volume (Mean ± SEM; \**p* < 0.05, \*\**p* < 0.01, one-way ANOVA). **(J)** CSF flow assessed by cisterna magna injection of high molecular weight dextran (70 kDa, green). Fluorescence stereo microscope images displaying the spatial distribution of the glymphatic system (green) in the brains (*top row*), and corresponding whole brain fluorescence imaging system scans of the same brains(*bottom row*), scale bars: 2 mm.

To visualize DDR2 distribution and its vascular association in vivo, we developed a fluorescent single-domain antibody (sdAb) probe targeting DDR2 (1A12-mCherry). The probe specifically labeled DDR2-positive cells (fig. S7, A to C), with signal intensity elevated in AD mice and further increased upon DDR2 overexpression (Fig. 3, C and D). Following the experimental timeline outlined in Fig. 3E, sequential injection of 1A12-mCherry and Dextran-FITC revealed reduced vascular perfusion in AD brains, a deficit that was further exacerbated by astrocyte-specific DDR2 overexpression (Fig. 3F). Two-photon laser scan microscope (TPLSM) imaging showed perivascular and parenchymal DDR2 accumulation in overexpressing brains, with AD mice displaying fragmented capillary networks and irregular vessel morphology, deficits aggravated by DDR2 overexpression (Fig. 3G and fig. S7D). Notably, astrocyte-specific DDR2 overexpression in wild-type mice was sufficient to reduce cerebral perfusion (fig. S8, A to C), indicating a direct effect on vascular function.

Vascular integrity assessment using size-fractionated dextran tracers revealed marked leakage of small molecules (4 kDa) into the parenchyma of AD mice, which was exacerbated by DDR2 overexpression, with near complete loss of small vessel retention and diffuse parenchymal saturation (Fig. 3H and fig. S9A). Mechanistically, DDR2 overexpression further reduced pericyte coverage, tight junction protein expression (ZO1, Claudin 5, Occludin), and Collagen IV, a critical structural component of the vascular basement membrane essential for BBB integrity (*23, 24*) in AD brains (fig. S9, B to H), consistent with progressive BBB breakdown.

Rodent glymphatic studies demonstrate CSF flow from ventricles into brain parenchyma along periarterial spaces (*25, 26*). Given DDR2 expression in CPECs, we examined ventricular morphology and CSF dynamics. Magnetic resonance imaging (MRI) volumetric reconstruction revealed progressive ventricular enlargement with increasing DDR2 expression (Fig. 3I). Cisterna magna tracer injection showed that CSF flow and parenchymal penetration were markedly impaired in AD mice and further diminished by DDR2 overexpression (Fig. 3J and fig. S10, A and B), indicating that DDR2 disrupts both vascular perfusion and CSF transport.

Collectively, these data demonstrate that astrocytic DDR2 overexpression impairs CSF dynamics and vascular integrity in AD mice, driving ventricular enlargement, reduced parenchymal tracer penetration, and basement membrane deterioration. These findings establish DDR2 as a key mediator of brain fluid clearance failure and neurovascular dysfunction in AD.

### Structural basis of HL2-mediated DDR2 recognition and lysosomal degradation

Having established that HL2 recognizes the native conformation of the DDR2 extracellular domain, we next assessed its binding characteristics and functional potential. Surface plasmon resonance (SPR) revealed that HL2 binds DDR2 with high affinity, exhibiting a *K_D_* of 0.519 nM, a *K_on_* of 5.68 × 10⁵ M⁻¹S⁻¹, and a *K_off_* of 2.95 × 10⁻⁴ S⁻¹ (Fig. 4A). To elucidate the structural basis of this interaction, we next reconstituted a binary complex consisting of the HL2 Fab fragment and the discoidin domain of DDR2 (DDR2-DS) (fig. S11) and determined its crystal structure at a resolution of 2.25 Å (Fig. 4B and tab. S2). As previously reported (*27, 28*), DDR2-DS adopts an eight-stranded (β1-β8) β-barrel fold composed of two antiparallel β-sheets. Structural superposition revealed high conformational similarity between HL2-bound DDR2-DS and its collagen-bound state (fig. S12), with a Cα root-mean-square deviation (RMSD) of 0.295 Å, indicating that HL2 binding does not induce substantial structural rearrangements. HL2 targets the “waist” region of the DDR2-DS β-barrel by recognizing one face of the three-stranded (β8-β3-β6) β-sheet (Fig. 4C). Comparison with the published DDR2-DS-collagen complex confirmed that the HL2 epitope at the DDR2-DS “waist” is entirely distinct from the collagen-binding site located at the top of DS domain (Fig. 4C). This structural observation suggests that HL2 does not directly compete with collagen for DDR2 binding, which is further supported by a pull-down assay showing that DDR2-DS retains collagen-binding capacity in the presence of HL2 (Fig. 4D and E).

**Fig. 4.**
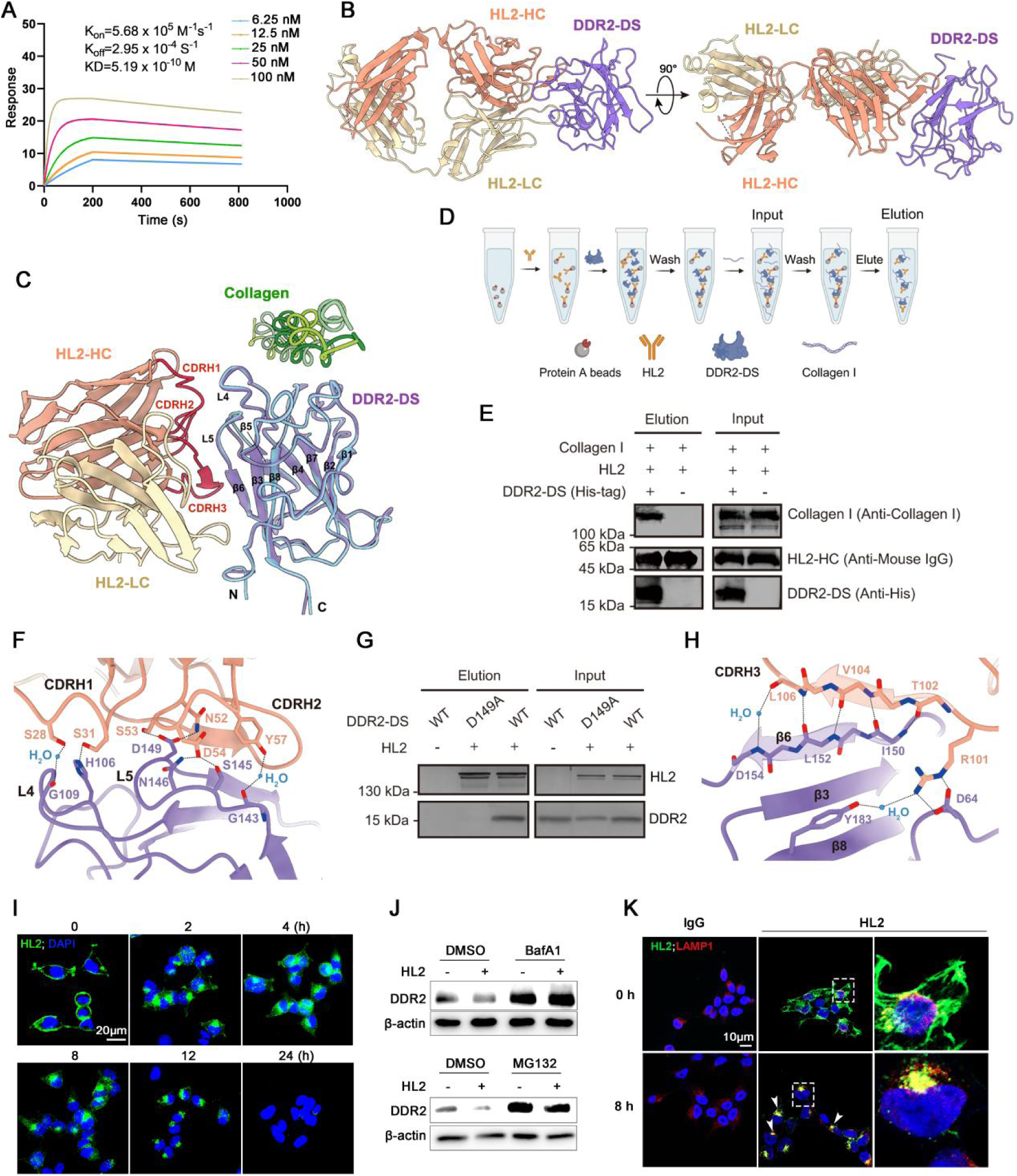
Characterization of the high- affinity anti- DDR2 monoclonal antibody HL2. **(A)** Representative sensorgrams showing HL2 binding to immobilized human DDR2 at increasing concentrations (0.625-10 nM) from the SPR analysis. The association (*K_on_*) and dissociation (*K_off_*) phases are fitted to a 1:1 binding model. **(B)** Overall structure of the DDR2-DS–HL2-Fab complex. HL2-Fab heavy chain (HL2-HC), light chain (HL2-LC) and DDR2-DS are shown as ribbons and coloured in orange, yellow and purple, respectively. **(C)** Structural superposition of the DDR2-DS–HL2-Fab complex with the DDR2-DS–collagen complex (PDB: 2WUH). Constant regions of HL2-Fab are omitted for clarity. DDR2-DS and collagen from the DDR2-DS–collagen complex are coloured in blue and green, respectively. The three CDRs (CDRH1–3) are highlighted in red and labeled. Secondary structural elements (β1–β8, L4 and L5) of DDR2-DS are indicated as previously described (*27*). **(D)** Schematic diagram of the collagen pull-down assay. **(E)** HL2 antibody-bound DDR2-DS retains collagen-binding activity. All pull-down experiments were repeated two times. **(F)** Polar interactions between DDR2-DS and HL2 at the CDRH1 and CDRH2 regions. Critical residues are shown as sticks and labeled. Water molecules involved in these interactions are depicted as blue spheres, and polar interactions are indicated with dashed lines. **(G)** The DDR2-DS D149A mutant abrogates binding to HL2. **(H)** Polar interactions between DDR2-DS and HL2 at the CDRH3 region. To highlight backbone-mediated contacts between CDRH3 and DDR2-DS β6, only the backbone atoms are shown as sticks, with the two corresponding β-strands superimposed as translucent ribbons. **(I)** Representative fluorescent images from HL2 internalization using a fluorescent secondary antibody (green) at different time points, scale bars: 20 μm. **(J)** Representative western blot of DDR2 and β-actin from 293T-DDR2 cells pretreated with the lysosomal inhibitor BafA1 (100 nM) or the proteasomal inhibitor MG132 (10 µM) for 1 hour, followed by incubation with HL2 (10 µg/ml) for 24 hours. **(K)** Representative fluorescent images of HL2 (green) and LAMP1 (red) in 293T-DDR2 cells incubated with HL2 for 8 h, scale bars: 10 μm.

Detailed structural analysis revealed that interactions between HL2 and DDR2-DS are predominantly mediated by the heavy chain, with all three complementarity-determining regions (CDRs) contributing to DDR2-DS binding (Fig. 4C). Specifically, CDRH1 primarily engages the L4 loop of DDR2-DS (Fig. 4F). Ser28 forms a water-mediated hydrogen bond with the backbone carbonyl oxygen of Gly109^D^ (superscript D denotes DDR2-DS residues), while Ser31 directly interacts with His106^D^ via a hydrogen bond. CDRH2 residues (Asn52–Asp54 and Tyr57) establish a hydrogen bond network with the L5 loop of DDR2-DS, involving residues Asp149^D^, Ser145^D^, Asn146^D^, and Gly143^D^ (Fig. 4F). Mutation of Asp149^D^ to alanine (D149A) abolished HL2 binding (Fig. 4G), underscoring the critical role of CDRH2 in DDR2 recognition. Notably, residues Thr102–Leu106 within CDRH3 fold into a short β-strand that packs in parallel with the β6 strand of DDR2-DS (Ile150^D^-Asp154^D^), stabilized by an extensive backbone-mediated hydrogen bond network (Fig. 4H). Additionally, Arg101 in CDRH3 further reinforces the interface via a salt bridge with Asp64^D^ and a water-mediated hydrogen bond with Tyr183^D^. Beyond these polar contacts, HL2 engages DDR2-DS via hydrophobic interactions. CDRH3 residues (Val103, Leu105, and Leu111) together with CDRL1 residues (Tyr26 and Ala27) form a hydrophobic cluster with Leu99^D^, Ile150^D^, Leu152^D^, and Tyr183^D^ (fig. S13A), while Tyr57, Val104, and Leu106 collectively cradle Phe151^D^ in β6 strand (fig. S13B). Collectively, these structural observations delineate the epitope recognized by HL2 and confirm that it is entirely non-overlapping with the collagen-binding site on DDR2.

Antibody-mediated receptor internalization is a well-established mechanism by which therapeutic antibodies modulate cell surface signaling. Having established that HL2 binds DDR2 with high affinity at a non-competitive epitope, we next assessed whether HL2 modulates DDR2 levels through this mechanism. We incubated 293T-DDR2 cells with HL2 under different conditions. Internalization was minimal at 4°C but robust at 37°C, detectable within 0.5 hours and exceeding 60% by 8 hours (fig. S14). Continuous incubation at 37°C for 24 hours resulted in near-complete loss of surface signal, indicating over 90% internalization (Fig. 4I). HL2 treatment significantly reduced DDR2 protein levels, an effect blocked by the lysosomal inhibitor BafA1 but not by the proteasomal inhibitor MG132 (Fig. 4J). Consistently, internalized HL2 colocalized with the lysosomal-associated membrane protein 1(LAMP1) (Fig. 4K), indicating that HL2 directs DDR2 to lysosomal degradation.

### AAV-mediated CNS delivery of HL2 attenuates AD pathology and cognitive decline

To evaluate the therapeutic potential of HL2 in vivo, we first administered purified HL2 protein systemically to APP/PS1 mice. However, brain levels of the antibody remained undetectable (data not shown), consistent with the less than 1% brain penetration reported for therapeutic Aβ monoclonal antibodies. To overcome this limitation, we delivered HL2 via a single systemic dose of AAV-PHP.eB, a BBB-permeable serotype with high tropism for the CNS (*29, 30*), encoding the antibody to 10-month-old APP/PS1 mice and monitored outcomes over 3 months following the experimental timeline outlined in Fig. 5A. Circulating HL2 levels in peripheral blood remained negligible at one- and two-month post administration (fig. S15). In vivo imaging revealed no change in cerebral DDR2 signal at 1 month after administrated, but a marked reduction by 2 months following HL2 treatment (Fig. 5, B to D). Behaviorally, HL2-treated AD mice showed improved spatial learning and memory in the Morris water maze, with a 50% reduction in escape latency and a 33% increase in target quadrant occupancy (Fig. 5E). Concurrently, AV45-PET imaging revealed substantially reduced Aβ deposition (Fig. 5F). At 3 months post treatment, HL2 was robustly expressed in the brain, with over 80% of cells positive (Fig. 5G). Immunofluorescence confirmed widespread reduction of DDR2 expression in cortex, hippocampus, and choroid plexus (Fig. 5H), accompanied by downregulation of Collagen X (Fig. 5I). HL2 treatment also rescued neuronal loss (Fig. 5J) and reduced both Aβ plaque burden and BACE1 expression (Fig. 5, K and L).

**Fig. 5.**
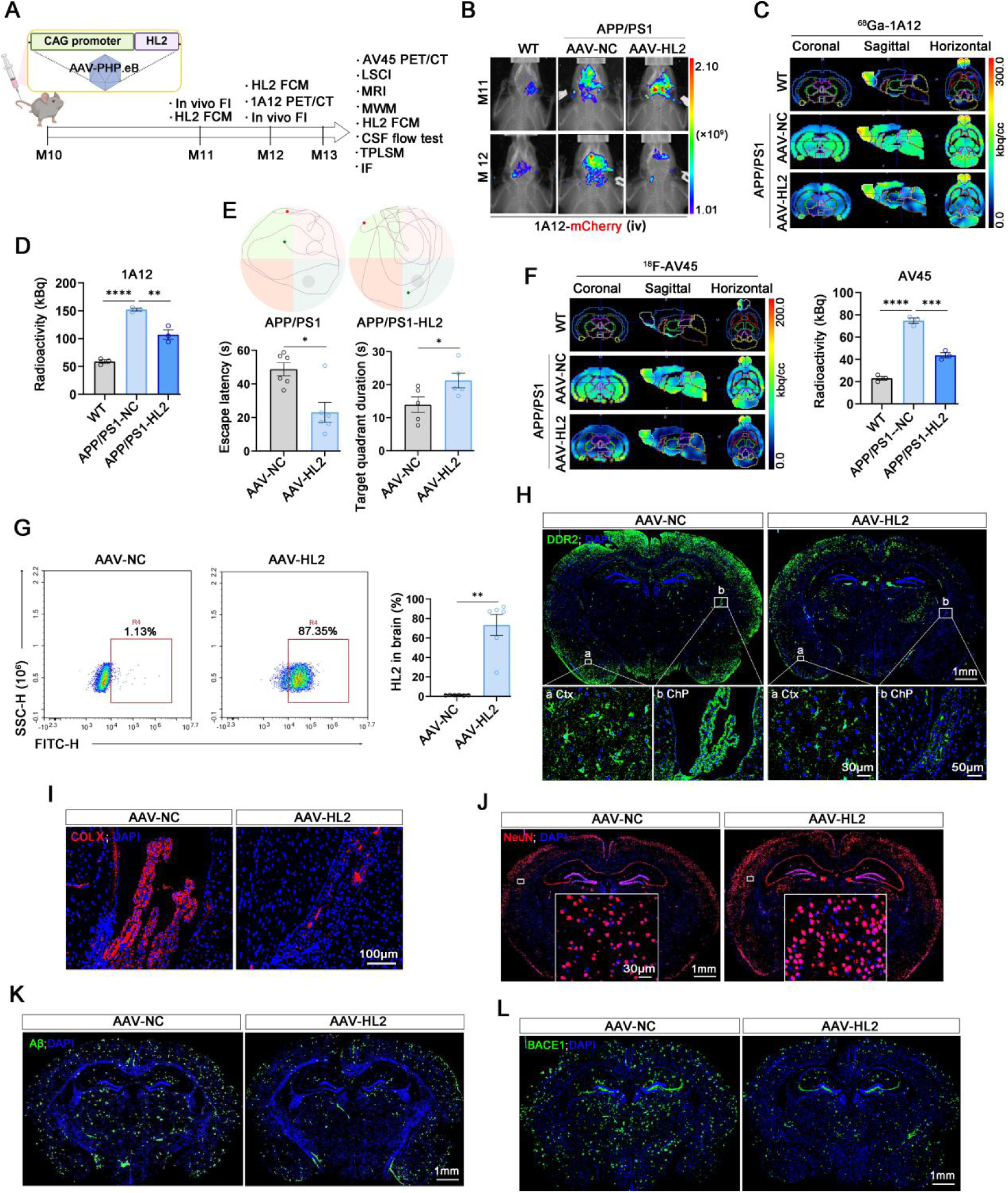
Effects of HL2 on pathology and cognitive deficits in APP/PS1 mice. **(A)** Experimental timeline: 10 month old APP/PS1 mice received a single systemic injection of AAV-PHP.eB packaged HL2. Longitudinal assessments included *in vivo* imaging at 1 and 2 months, behavioral testing at 3 months, and terminal tissue collection for biochemical and histopathological analysis. **(B)** *In vivo* whole brain fluorescence imaging using a DDR2 targeting sdAb probe 1A12-mCherry. **(C)** Representative multi planar PET/CT sections depicting regional tracer uptake: coronal (*left*), sagittal (*middle*), and horizontal (*right*) views *in vivo* PET/CT imaging using a DDR2-targeting probe (1A12). Color scale indicates standardized uptake intensity. **(D)** Quantification of brain DDR2 signal intensity. **(E)** Representative motion tracks of APP/PS1 and APP/PS1 transgenic mice treated with HL2 in morris water maze performance (*top*); Escape latency during training days (*bottom left*) and the target quadrant duration during the trial (*bottom right*). Data were shown as mean ± SEM and paired *t*-tests analysis was used for data analysis (\**p* < 0.05). **(F)** *In vivo* AV45-PET imaging of amyloid burden after HL2 treatment (*left*), and quantification of whole-brain AV45 uptake (*right*). **(G)** Representative flow cytometry plots showing the HL2 expression in suspensions from whole brain homogenates at 3 months post treatment (*left*), and quantification of the percentage of HL2 positive cells in total live cells. Data are presented as mean ± SEM, \*\**p* < 0.01, paired *t*-tests. (H) Down-regulation of DDR2 after HL2 treatment. **(H)** Representative immunofluorescence images of DDR2 (red) and DAPI (blue) in cortex and CPECs. Scale bars: 100 μm (cortex, hippocampus). Scale bar: 1 mm for original images, 30 μm for enlarged images in cortex, and 50 μm for enlarged images in CPECs. **(I)** Representative staining for Collagen X (red) in APP/PS1 mice after HL2 treatment. Scale bar: 100 μm. Representative staining for NeuN (red) **(J),** Aβ (green) **(K)** and BACE1 (green) **(L)** in the coronal section of a mouse brain. Scale bar: 1 mm for original images, 30 μm for enlarged images.

Together, these data demonstrate that a single systemic administration of AAV-delivered HL2 achieves sustained brain expression, suppresses DDR2 and downstream pathological markers, including Aβ deposition, BACE1, and Collagen X, while preserving neurons and improving cognitive function in AD mice. These findings establish DDR2 as a viable therapeutic target for modulating core disease pathways in AD.

### HL2-mediated DDR2 downregulation restores cerebral perfusion, vascular integrity, and CSF dynamics

Given the observed improvements in AD pathology and cognition following HL2 treatment, we next examined its effects on the cerebrovasculature and glymphatic system. LSCI revealed that HL2 treatment increased cerebral perfusion by approximately 16% in AD mice (Fig. 6A). Given that ^18^F-FDG uptake correlates positively with hemodynamics and is reduced in early AD (*31, 32*), we next assessed glucose metabolism. Consistent with clinical observations in early AD (*33, 34*), HL2-mediated DDR2 downregulation significantly reversed the regional hypometabolism observed in the hippocampus, thalamus, and hypothalamus of AD mice (Fig. 6B).

**Fig. 6.**
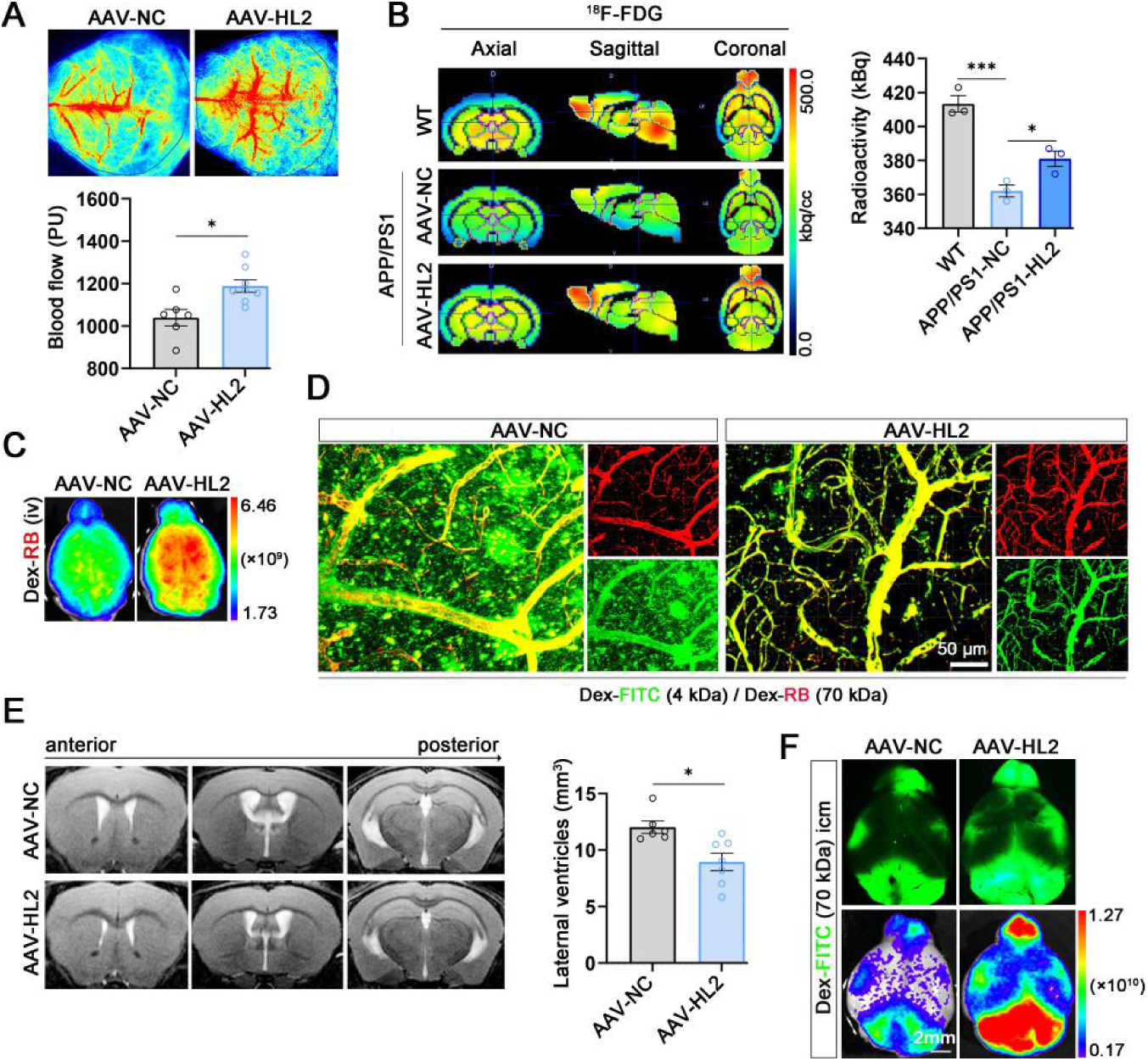
Effects of HL2 on the cerebral hemodynamics and CSF dynamics in AD Mice. **(A)** Representative pseudocolor perfusion maps of the cortical surface by LSCI (*top*). Quantification of relative perfusion units (*bottom*). Data are presented as mean ± SEM, \**p* < 0.05, paired *t*-tests. **(B)** Cerebral glucose metabolism measured by ¹⁸F-FDG PET/CT. Left: Representative coronal (*left*), sagittal (*middle*), and horizontal (*right*) PET images superimposed on CT templates, showing various brain ROIs. Right: Quantification of ¹⁸F-FDG standardized uptake value within the combined ROIs. Data are presented as mean ± SEM, \**p* < 0.05, \*\*\**p* < 0.001, one-way ANOVA. **(C)** Whole brain fluorescence imaging of vascular perfusion with Dextran-FITC (70 kDa) in APP/PS1 mice after HL2 treated. **(D)** Vascular integrity assessed by dual tracer dextran perfusion: representative two-photon microscopy images of vascular leakage after injection of dextran-FITC (4 kDa, green) and dextran-RB (70 kDa, red), scale bars: 50 μm. **(E)** Ventricular morphology was evaluated using MRI-based volumetric reconstruction. Left: Representative images of lateral ventricles across groups. Right: Quantitative analysis of lateral ventricular volume (Mean ± SEM; \**p* < 0.05, paired *t*-tests). **(F)** CSF flow was examined via cisterna magna injection of high molecular weight dextran (70 kDa, green). Top row: Fluorescence stereomicroscope images illustrating glymphatic pathway distribution (green) across brain sections. Bottom row: Corresponding whole-brain fluorescence scans of the same specimens. Scale bars: 2 mm.

Vascular integrity assays using dextran tracers demonstrated enhanced whole brain vascular signal in HL2 treated mice compared to AD controls (Fig. 6C). At 30 min post injection, TPLSM revealed reduced vascular leakage, as evidenced by diminished parenchymal extravasation of 4 kDa dextran and preserved intravascular signal in small vessels (Fig. 6D and fig. S16A). Immunofluorescence and qPCR analyses revealed that HL2 treatment upregulated pericyte coverage (PDGFRβ), basement membrane components (Collagen IV), and tight junction proteins (Occludin, ZO1) (fig. S16, B to E). Consistent with restored vascular integrity, HL2 treatment reduced lateral ventricular volume by 25% (Fig. 6E) and mitigated CSF flow impairment in AD mice (Fig. 6F), indicating improved glymphatic function.

Collectively, these findings demonstrate that HL2-mediated DDR2 downregulation enhances cerebral perfusion, restores glucose metabolism, improves vascular barrier integrity, and normalizes CSF dynamics, underscoring its multifaceted therapeutic potential in ameliorating neurovascular dysfunction and CSF dynamics associated with AD.

## Discussion

Here, we present a comprehensive spatial atlas of DDR2 expression in the AD brain, identifying it as a previously unrecognized nodal driver of multi-compartmental failure. Our analysis reveals marked upregulation of DDR2 in three functionally distinct cell populations: PVFs, reactive astrocytes, and CPECs. Hypoxia is a known inducer of DDR2 expression (*35, 36*). Given that reduced CBF in AD leads to tissue hypoxia, this may partially account for the elevated DDR2 levels observed in the disease. The convergence of DDR2-driven pathology across these cell types implicates this receptor as a master regulator of vascular and glymphatic dysfunction in AD.

The increase of DDR2 in PVFs is consistent with its upregulation in aged meningeal fibroblasts (*37*), a cell population now recognized as a principal origin of PVFs (*38*), and aligns with the receptor’s well-established role in pro-fibrotic signaling. Despite constituting less than 1% of total brain cells, PVFs ensheathe the major cerebral vessels that control blood inflow and outflow. Mirroring its function in IPF, where DDR2 cooperates with TGF-β and collagen to sustain fibrosis (*12, 39*), elevated DDR2 in AD brain PVFs likely drives pathological perivascular fibrosis. Supporting this, we observed abundant Collagen Ⅰ deposition surrounding DDR2-positive vasculature in human AD tissue, suggesting that fibrotic remodeling might impair flow through the brain’s primary conduits for blood and CSF.

In the glial compartment, the burden of *DDR2^+^* reactive astrocytes positively correlates with Braak stage in individuals with AD, and these cells exhibit a neurotoxic phenotype. In mice, astrocyte specific DDR2 overexpression recapitulated this pathology, impairing both CBF and glymphatic influx, while concurrently upregulating BACE1, the rate-limiting enzyme for Aβ production, and accelerating Aβ deposition. This positions DDR2 as an upstream regulator of astrocyte-driven Aβ synthesis. Critically, while BACE1 itself has long been pursued as a therapeutic target, direct pharmacological enzymatic inhibition has proven challenging due to broad off-target toxicity (*40*). DDR2 thus represents an upstream, cell-type-specific node for modulating the amyloid cascade via astrocytic BACE1. By acting upstream to specifically drive astrocytic *Bace1* expression, DDR2 may represent a more tractable node for intervention, enabling cell-type-specific modulation of the amyloid cascade. Beyond parenchymal accumulation, this Aβ overproduction directly compromised cerebrovascular integrity, evidenced by reduced CBF and increased vascular leakage. This pathology aligns with cerebral amyloid angiopathy, wherein perivascular Aβ deposits induce smooth muscle degeneration, basement membrane thickening, and perivascular fibrosis (*41*). Thus, the Aβ overload driven by astrocytic DDR2 directly contributes to the observed vascular pathology, establishing a mechanistic link between glial reactivity and both hemodynamic compromise and BBB disruption.

Beyond PVFs and astrocytes, DDR2 may also compromise CSF dynamics through its action on CPECs, the specialized epithelium responsible for CSF secretion. In CPECs, DDR2 was markedly upregulated and co-localized with Collagen X, a non-fibrillar collagen known to activate DDR2 (*20*). Treatment with a DDR2-targeting antibody (HL2) reduced both proteins, revealing that DDR2 activity sustains Collagen X expression. These findings reframe the enigmatic phenomenon of choroid plexus calcification. Collagen X is not merely a matrix constituent but a well-established initiator of calcification in hypertrophic cartilage (*42*). Our data thus raise the possibility that DDR2, by driving Collagen X synthesis, actively promotes epithelial calcification. While choroid plexus calcification has historically been viewed as an inert byproduct of inflammation (*43*), our findings implicate a self-sustaining Collagen X-DDR2-Collagen X cascade in this process, which may in turn disrupt CSF homeostasis. Formal validation of this hypothesis will require CPEC-specific genetic models. Targeting DDR2 may therefore offer a strategy to preserve choroid plexus function and restore brain environment stability.

Our structural study reveals that the monoclonal antibody HL2 binds specifically to the “waist” region of the DDR2 DS domain, an epitope distinct from the collagen-binding site located at the top of DS β-barrel fold (*28*), as further validated by competitive binding assays. This non-overlapping epitope enables HL2 to efficiently engage and internalize DDR2 regardless of collagen occupancy, effectively depleting the pathogenic receptor pool within collagen-enriched fibrotic microenvironments. This mechanism contrasts with that of small-molecule DDR2 kinase inhibitors, which primarily block collagen-induced activation (*44, 45*).

To achieve efficient and sustained CNS delivery of HL2, we employed an AAV-mediated expression strategy. AAV-mediated delivery of the antibody resulted in high-titer expression confined to the <u>CNS,</u> with minimal to undetectable levels in peripheral tissues. This restricted expression profile suggests a reduced risk of off-target effects on skeletal or reproductive tissues, consistent with the predominant phenotypes, skeletal abnormalities and infertility, observed in DDR2 knockout models (*46*). The sustained CNS expression of this AAV-delivered antibody is anticipated to exhibit a favorable safety profile, supported by low basal DDR2 levels in the normal adult brain and a knockout phenotype primarily restricted to skeletal abnormalities. As an alternative to AAV-based delivery, recent advances in antibody transport across the BBB offer a complementary strategy for clinical translation. Bispecific antibodies engineered to harness receptor-mediated transcytosis, most prominently via the transferrin receptor (TfR), have achieved robust CNS penetration (*47*).

The clinical success of Aβ and tau immunotherapies in AD has been inextricably linked to cognate PET tracers, now indispensable for patient enrollment, target engagement, and treatment monitoring (*48*). This paradigm reflects a broader reality in neurology: tissue biopsy is rarely feasible, and targeted therapies increasingly depend on companion diagnostics. Building on this principle, we developed a DDR2-targeted sdAb PET tracer, which was validated in both animal models a first-in-human IPF study, where it successfully visualized active fibroblast-rich foci (https://medrxiv.org/cgi/content/short/2025.11.26.25341068v1). Given that astrocytes vastly outnumber fibroblasts in the AD brain, we hypothesize that the DDR2-PET signal would derive predominantly from DDR2-positive reactive astrocytes. Supporting its applicability in this context, a fluorescently labeled DDR2 sdAb confirmed BBB penetrance and CNS target engagement. Astrocyte activation is an early event in AD, and DDR2 is enriched in neurotoxic astrocytes. DDR2-PET therefore offers a long-needed tool to visualize this pathogenic neuroinflammatory response in vivo. It holds potential to detect AD-related changes prior to detectable Aβ accumulation on conventional PET, a hypothesis that awaits clinical validation. Thus, paralleling the role of Aβ and tau PET for their respective immunotherapies, DDR2-PET could serve as a vital companion diagnostic for DDR2-targeted therapies.

Accumulating evidence of cognitive decline in IPF patients (*49, 50*) aligns with our identification of DDR2 as a shared driver in both IPF and AD. In the brain, perivascular fibrosis compromises CBF and CSF circulation, supporting the emerging concept of AD as a CNS fibrotic disease (*51*). We show that DDR2 co-expresses with fibrillar collagens (Collagen I/III) in PVFs and with non-fibrillar Collagen X in CPECs, revealing cell-type-specific collagen programs. Although the collagen repertoire of DDR2^+^ astrocytes remains to be defined, perivascular astrocytes adjacent to the PVS may be activated via DDR2 engagement with fibrillar collagens in the fibrotic niche. Together, these cell-type-specific mechanisms converge to disrupt vascular and glymphatic function in the AD brain. These findings provide a rationale for translating fibrosis-directed strategies from peripheral diseases to AD. Supporting this, our unpublished data show that HL2 also exerts antifibrotic efficacy in IPF models (unpublished data), underscoring the potential of targeting DDR2 for lung-brain comorbidities.

This study uncovers DDR2 as a central mechanistic hub that integrates vascular, glymphatic, and choroid plexus dysfunction in AD. By coupling a CNS-penetrant, AAV-delivered antibody for durable target ablation with a companion PET tracer for non-invasive monitoring, we establish a first-in-class theranostic platform. This integrated approach enables targeted restoration of cerebral energy infrastructure, offering a previously inaccessible strategy for correcting multi-compartmental failure in AD.

## Materials and Methods

### scRNAseq analysis

FASTQ files were obtained from the Gene Expression Omnibus (GEO) and aligned to the hg38 reference genome. Cell type identification was performed with Cell Ranger (v6.0.0) for barcode assignment and unique molecular identifier (UMI) counting. The raw UMI count matrix (genes × cells) was then processed through the SCANPY pipeline to perform quality control and downstream analyses (*52*). Cells with >10% mitochondrial gene content or fewer than 100 detected genes were excluded. Genes detected in fewer than 50 cells were filtered out. For each cell, raw counts were normalized to total library size and log-transformed after adding a pseudocount. Highly variable genes were selected with pp.highly_variable_genes function and subsequently used for cell clustering. Principal component analysis (PCA) was performed on highly variable genes, and batch effects were corrected using Harmony (*53*). A neighborhood graph was computed using pp.neighbors, followed by UMAP dimensionality reduction via tl.umap. Cluster marker genes were identified by ranking differentially expressed genes in each cluster using tl.rank_genes_groups. For pseudotime trajectory analysis, Monocle2 was employed with default settings to infer gene expression dynamics in astrocytes (*54*).

### Purification and affinity assay of HL2-mFc

A chicken egg yolk-derived polyclonal antibody (IgY clone HL2) was raised against the purified extracellular domain of human DDR2. To generate a chimeric HL2 antibody (HL2-mFc) suitable for mammalian immunoassays, a synthetic heavy-chain construct consisting of the HL2 variable heavy (VH) region fused to the mouse IgG2a Fc region, and a corresponding light-chain construct consisting of the HL2 variable light (VL) region fused to the mouse IgG2a constant light (CL) region, were co-expressed in 293-F cells. The recombinant antibody was secreted into the culture supernatant and purified using Protein A affinity chromatography (GenScript). An international patent application has been filed for this antibody (PCT/CN2026/074630).

Binding analysis of HL2-mFc *in vitro*: To evaluate HL2-mFc specificity, DDR2 was transiently overexpressed in HEK293T cells. Both parental HEK293T and DDR2-transfected cells were fixed on coverslips, incubated with HL2-mFc, and subsequently with an Alexa Fluor 647-conjugated anti-mouse IgG secondary antibody (1:2,000 dilution; Abcam, ab150123). Binding was assessed by fluorescence microscopy and quantified by flow cytometry using a NovoCyte instrument (ACEA Biosciences).

*In vivo* validation in mouse brain: For *in vivo* detection of DDR2, adult C57BL/6 mice received intracranial injections of an AAV-PHP.eb vector encoding full-length DDR2 under a constitutive promoter. Three weeks post-injection, mice were transcardially perfused with PBS followed by 4% paraformaldehyde. Brains were post-fixed overnight, sectioned, and subjected to immunofluorescence staining as described below.

### Immunofluorescence staining

Frozen tissue sections were prepared by embedding brain or spinal cord tissues in optimal cutting temperature (OCT) compound, followed by sectioning into 10 μm thick slices and fixation for further use. Sections were washed three times with PBS, blocked with 5 % BSA for 1 h, and incubated with primary antibody overnight at 4 ℃. After washing in PBS, sections were incubated with species-appropriate Alexa secondary antibody for 2h at room temperature. The sections were then stained with DAPI, and the slides were scanned using the Pannoramic MIDI slide scanner (3DHISTECH). The primary antibodies used in this study were as follows: HL2-mFc for DDR2 (1 μg per sample), anti-Aβ (1:500; BioLegend, SIG-39220), anti-GFAP (1:200; abcam, ab16997), anti-NeuN (1:500; Millipore, ABN78), anti-CD31 (1:1000; Cell Signaling Technology, no. 3528S), anti-DCN (1:200; abcam, ab175404), anti-Collagen X (1:200; abcam, ab49945 for human and ab260040 for mouse), anti-S100A10 (1:100; Thermo Fisher, MA5-24769), anti-BACE1 (1:100; Cell Signaling Technology, no. 5606), anti-Collagen Ⅳ (1:200; abcam, ab6586), anti-PDGFRβ (1:100; Cell Signaling Technology, no. 3169), anti-Occludin (1:100; Cell Signaling Technology, no. 91131).

### Affinity assay of CBP-FITC

Isolation of mouse primary lung fibroblasts (mPLFs): Mouse lung tissues were minced into small pieces and incubated in Dulbecco’s Modified Eagle’s Medium (DMEM, GIBICO, Life Technologies Corporation) containing collagenase II (2 mg/ml), trypsin (2.5 mg/ml), DNase I (2 mg/ml), penicillin (100 U/ml), and streptomycin (100 μg/ml) for 12 h at 37°C. The tissues were then washed with Dulbecco’s phosphate-buffered saline (DPBS). Cells were collected from the supernatant after filtering out residual tissue fragments using a 70 μm cell strainer. The isolated cells were cultured in DMEM supplemented with 10% heat-inactivated fetal bovine serum (FBS, Gibco, no.10099141). High-purity fibroblast populations were obtained after at least three passages. 3×10^5^ mPLFs at 50% confluence were plated in a 6-well plate and treated with 10 ng/ml TGF-β1 and *Col1a1* siRNA for 48h.The expression of *Col1a1* was assessed by quantitative PCR, and the staining for CBP was illustrated as follows: Cell supernatant was removed after 48 hours, and then cells were fixed with 4% paraformaldehyde for 15 minutes, followed by incubated with 4 µg/ml CBP-FITC in the dark on a shaker at 150 rpm/min for 1 hour at room temperature. Following washed by PBS 3 times, nuclei were stained with DAPI for 5 minutes. Cells were imaged using a ZEISS LSM900 confocal fluorescence microscope.

### Preparation of ^68^Ga-NOTA-1A12 and PET/CT imaging

DDR2 specific single domain antibody (sdAb), designated 1A12, was generated, purified, and conjugated with ^68^Ga following established protocols from our recent study (https://medrxiv.org/cgi/content/short/2025.11.26.25341068v1). Mice PET/CT imaging was performed on a MadicLab PSA094 PET/CT scanner (Shandong Madic Technology). The image data was collected 30 min after the injection of 7.4 MBq of ^68^Ga-1A12, ^18^F-AV45, or ^18^F-FDG while the mice were anesthetized using 1.5% isoflurane inhalation anesthesia.

### Image analysis

PET images were reconstructed in 3D OSEM with the CT scan data for the attenuation correction and anatomical references. All data was processed and analyzed with the PMOD software (version 3.301, PMOD Technologies). The images were displayed as coronal, sagittal, and transverse sections, excluding the activities outside the cerebral. Using the PMOD PFUS module, the PET/CT images of the mouse brain were spatially normalized and fitted to the template of magnetic resonance imaging (MRI) of the mouse brain included in the PMOD software, and scaled to the Paxinos and Franklin coordinate system. The average activity per volume unit (kBq/cc) was subsequently corrected for the injected radioactivity and mouse weight, and expressed in the unit of standardized uptake value (SUV). Based on the fitted mouse MRI template, eight volumes of interest (VOIs) were selected for further evaluation, namely the striatum (red), cortex (blue), hippocampus (dark green), hypothalamus (pale green), thalamus (green), amygdala (yellow), olfactory bulb (white), and midbrain (pale purple). The images and region of interest (ROI) for mouse spinal cord were analyzed using RadiAnt software.

For quantitative analysis, PET images were reconstructed using a three-dimensional ordered subset expectation maximization (3D OSEM) algorithm, with CT data applied for attenuation correction and anatomical reference. All imaging data were processed and analyzed using PMOD software (version 3.301; PMOD Technologies Ltd., Zurich, Switzerland). Images were displayed in coronal, sagittal, and transverse planes. The average radioactivity concentration (kBq/cc) within each region was normalized to the injected dose and body weight of the mouse, and expressed as the standardized uptake value (SUV).

### Animal models

All animal protocols used in this study conformed to the requirements of the Animal Welfare Act and were approved by the Animal Care and Use Committee of Guangzhou Medical University (Approval No. 20230411).

AD mice: Male APPswe/PSEN1dE9 (APP/PS1) mutated transgenic mice with a C57BL/6J background were purchased from Jackson Laboratory guidelines (https://www.jax.org/strain/005864).

### Morris water maze (MWM) test

Spatial learning and memory were evaluated in a customized Morris water maze system consisting of a circular pool (120 cm diameter, 40 cm water depth) maintained at 22 ± 1°C. The protocol comprised three phases:

1. Habituation: Animals freely explored the pool for 60 s without platform.
2. Acquisition training: Over four consecutive days, mice underwent four daily trials to locate a submerged platform (10 cm diameter) in fixed quadrant. Each trial allowed 60 s exploration, with unsuccessful animals guided to platform and allowed 15 s reinforcement.
3. Probe testing: 24 h post-training, platform removal enabled 60 s free exploration. Spatial memory was quantified by target quadrant dwell time and platform-crossing frequency. Movement trajectories were tracked using EthoVision XT 15 software (Noldus).

### Real-time PCR

Total RNA was isolated from mice brain tissues using EZ-press RNA Purification Kit (EZBioscience, B0004D), followed by cDNA synthesis with PrimeScript™ Reverse Transcriptase (Takara, 2690S) according to manufacturer protocols. Quantitative real-time PCR was performed using 2 × SYBR Green qPCR Mix (EZBioscience, A0012). Primer sequences are as follows: (5’-3’):

Mouse *Ddr2*: Froward: GACTTGCGAACCCTACACTTT, Reverse: CGAACAAATCTGGCGACGAT
Mouse *Col4a1*: Froward: CCTGGCACAAAAGGGACGA, Reverse: ACGTGGCCGAGAATTTCACC
Mouse *ZO1*: Froward: GCCGCTAAGAGCACAGCAA, Reverse: GCCCTCCTTTTAACACATCAGA
Mouse *Bace1*: Froward: GGAGACCGACGAGGAATCG, Reverse: GCAAAGTTACTACTGCCCGTG
Mouse *Pdgfrb*: Froward: AGGAGTGATACCAGCTTTAGTCC, Reverse: CCGAGCAGGTCAGAACAAAGG
Mouse *Ocln*: Froward: TGAAAGTCCACCTCCTTACAGA, Reverse: CCGGATAAAAAGAGTACGCTGG
Mouse *β-actin*: Froward: GGCTGTATTCCCCTCCATCG, Reverse: CCAGTTGGTAACAATGCCATGT

Relative expression levels were normalized to β-actin using the 2^−ΔΔCt^ method.

### Western blotting

Tissues samples were lysed using ice-cold radioimmunoprecipitation assay lysis buffer (RIPA, Beyotime, P0013B) containing 1 % phenylmethanesulfonyl fluoride (PMSF, Beyotime, ST505) in a high-throughput tissue grinder (KeCheng, KC-48). After centrifuged, the supernatant was collected and the total protein concentration was detected by bicinchoninic acid protein assay kit (BCA, Biosharp, BS260). Protein was separated using SDS-PAGE and then transferred onto a poly (vinylidene fluoride) membrane (PVDF, Merck Millipore, ISEQ00005). After blocked in 5 % skim milk (Bioforxx, 1172GR100) at room temperature for 1 hour, the membranes were incubated with primary antibody at 4 ℃ overnight and then incubated with horseradish peroxidase (HRP)-conjugated secondary antibody at room temperature for 1 h. Finally, the specific protein band was visualized using an ultra-sensitive ECL chemiluminescent substrate (Biosharp, BL520A) in the GelView 6000 Pro Multifunctional Imaging Station (Biolight Biotechnology). The primary antibodies used in this study were as follows: anti-DDR2 (1:1000; Servicebio, GB112568), anti-DDR2 (1:1000, R&D, AF2538), anti-BACE1 (1:50; Cell Signaling Technology, no. 5606), anti-Collagen Ⅳ (1:1000; abcam, ab6586), anti-PDGFRβ (1:1000; Cell Signaling Technology, no. 3169), anti-Occludin (1:1000; Cell Signaling Technology, no. 91131), anti-ZO1 (1:1000; abcam, ab276131), anti-β-actin (1:1000; Yeasen, 30102ES60).

### Laser speckle contrast imaging (LSCI)

Mice were anesthetized by inhalation of 1% isoflurane in air via a mask and secured in a stereotaxic frame (RWD Life Sciences). The scalp was shaved, aseptically prepared with alternating povidone-iodine and 70% ethanol, and incised along the midline to expose the skull. The periosteum and fibrous tissue covering the skull were carefully removed by gentle abrasion with 3% hydrogen peroxide, followed by thorough rinsing and moistening with sterile saline to obtain a clear optical field. After surgical preparation and a 10-minute stabilization period, baseline speckle images were acquired for 20 seconds. Real-time changes in CBF were captured by a CCD camera. To ensure cross-group comparability, the same region of interest (ROI, indicated by a navy blue circle in the figures) was defined and analyzed for all animals.

### Preparation of mCherry-tagged 1A12 (1A12-mCherry)

To generate 1A12-mCherry, the coding sequence was cloned into a pCDNA3.1+ vector containing an N-terminal secretion signal. The construct was transfected into 293-F cells using polyethylenimine (PEI). Supernatant was harvested 120 hours post-transfection, filtered, and supplemented with 20 mM imidazole. Recombinant protein was purified via nickel-affinity chromatography, eluted with imidazole, and buffer-exchanged into PBS (pH 7.4). Purity was assessed by SDS-PAGE and Coomassie staining, and concentration was determined by A280. Binding affinity was validated by flow cytometry or fluorescence imaging. Purified protein was aliquoted and stored at −80°C for subsequent assays.

### *In vivo* fluorescence imaging

Mice were intravenously injected with 1A12-mCherry at a dose of 60 μg per mouse. After being shaved, they were anaesthetized with isoflurane and then the fluorescence images of lesion area were visualized with AniView600 Multi-Mode In Vivo Animal Imaging System with X-ray imaging module (Biolight Biotechnology). Wild-type C57BL/6J mice were used as negative controls ROI were performed using AniView 100 software (Biolight Biotechnology) with fluorescence radiance expressed as photons/s/cm^2^/sr.

### Two-photon microscopy imaging

Mice were then injected with 60 μg 1A12-mCherry, 1mg Dextran-FITC or Dextran-RB via the tail vein and sacrificed after 30 or 10 min post injection. Brains were harvested and promptly subjected to fluorescence imaging with an AniView600 Multi-Mode In Vivo Imaging System to survey global signal distribution. For detailed cellular or structural analysis, defined brain regions were further examined using a ZEISS LSM880 NLO Two-Photon Laser Scanning Microscope.

### MRI of the lateral ventricles in mice

Following induction with 2% isoflurane in a mixture of oxygen and air, mice were transferred to a head-fixed cradle integrated with a quadrature surface coil within a Bruker BioSpec 7.0 T/20 cm horizontal bore scanner (Bruker BioSpin). Anesthesia was maintained at 1.5% isoflurane. High-resolution T2-weighted images were obtained with a RARE-T2 sequence using the following parameters: TE = 52.0 ms, TR = 3600 ms, slice thickness = 0.7 mm, RARE factor = 8, FOV = 2.5 × 2.5 cm², echo spacing = 10.4 ms, averages = 2. Images were performed using Radient DICOM software (Medixant, Poland). The lateral ventricles were segmented semi automatically using a multi-step approach. First, a ventricle probability map was generated from the registered images using intensity thresholding and morphological operations. Manual corrections were then applied by a trained rater blinded to experimental conditions, using 3D Slicer, to ensure accuracy in regions of partial volume effect or low contrast.

### Cisterna magna injection and imaging

Mice were anaesthetized via intraperitoneal administration of 1% pentobarbital sodium (diluted in 0.9% saline). Following cervical hair removal and disinfection with 70% iodine, mice were placed on a stereotaxic apparatus to maintain the neck is protruding upward, and an ophthalmic solution was applied to prevent dry eyes. The skin from the neck was longitudinally incised, and muscles were retracted using hooks to expose the cisterna magna. The solutions were injected using a Hamilton syringe (10 μl; 1 μl min^−1^). The syringe was retained in place for 5 min to prevent back flow of the solution. Then the skin was sutured and mice were kept on a heating pad until fully awake. For evaluating CSF dynamics, or the mice were euthanized and transcardially perfused with PBS 1 hour after cisterna magna injection of FITC-dextran (70 kDa, 0.1mg in 10 μl), the whole brains were then placed on a Petri dish and imaged by Zeiss Axio Zoom V 16 stereofluorescence microscope for *Ex vivo* stereomicroscopy imaging. Subsequently, to further assess the diffusion profile of CSF within the brain parenchyma, the imaged brains were processed for cryosectioning and slide scanning. In a parallel experiment designed to test the penetration and distribution of smaller tracers, 4 kDa FITC-dextran (0.1mg in 10 μl) was injected into the cisterna magna, then the brains were harvested, cryosectioned, and subjected to high-resolution slide scanning to assess the parenchymal distribution and diffusion patterns of CSF tracers with varying molecular weights.

### Surface plasmon resonance (SPR) affinity measurement of HL2

The equilibrium dissociation constant of the HL2 monoclonal antibody for recombinant human DDR2 was determined by surface plasmon resonance on a Biacore 8K instrument (Cytiva). DDR2 was immobilized as the ligand on a CM5 sensor chip via amine coupling in sodium acetate (pH 4.0). A reference flow cell was prepared identically without ligand. Kinetic measurements were performed using a single-cycle kinetics format. Serially diluted HL2 (6.25 to 100 nM in HBS buffer) was injected sequentially over the chip surface at a flow rate of 30 µL/min, with a 200-second association and a 600-second dissociation phase for each concentration. The surface was regenerated between cycles with a 60-second pulse of 10 mM glycine-HCl (pH 1.5). Double-referenced sensorgrams were analyzed with Biacore Insight Evaluation Software (v3.0). Data were fitted to a 1:1 Langmuir binding model to derive the Association rate constant (Ka), Dissociation rate constant (Kd) and Equilibrium dissociation constant (K_D_).

### Protein expression and purification

For the expression of DDR2-DS domain, HEK293F cells were cultured in a humidified shaker with 5% CO_2_ and 55% humidity at 37 °C in SMM 293T-II (Sino Biological). The DNA fragment encoding the DS domain (residues 1-190) of DDR2 (UniProt: Q16832) was cloned into a pMLink vector with a C-terminal 8×His tag. The plasmid was transiently transfected into HEK293F cells using polyethylenimine (PEI; Polysciences). Five days after transfection, the conditioned medium was collected by centrifugation and dialysed to exchange into the binding buffer (8 mM Na_2_HPO_4_, 2 mM NaH_2_PO_4_, and 137 mM NaCl). The recombinant proteins were then isolated using Ni-NTA affinity purification and eluted with the binding buffer supplemented with 300 mM imidazole. The DDR2-DS domain was further purified using a Superdex 75 column (Cytiva) in the binding buffer. DDR2-DS mutants were expressed and purified similarly to WT DDR2-DS.

Papain digestion of IgG was performed to generate HL2-Fab. Briefly, papain (Sangon Biotech) was activated at 37°C for 15 min in the activation buffer containing 100 mM Tris-HCl, pH 8.0, 2 mM EDTA, and 10 mM L-cysteine. Then, 5 mg of IgG was incubated with the activated papain at an enzyme-to-antibody mass ratio of 1:100 in the same buffer (total volume 10 mL) at 37°C for 4-5 h. The reaction was terminated using 30 mM iodoacetamide at room temperature for 10 min. After termination, the buffer was exchanged to binding buffer using ultrafiltration, and the product was applied to Protein A beads (GenScript) to remove the Fc. The flow-through Fab was collected and further purified using a Superdex 75 column in the binding buffer.

### Crystallization and structure determination

Purified HL2-Fab was incubated with excess DDR2-DS for 30 min on ice, and the DDR2-DS/HL2-Fab complex was then isolated using a Superdex 75 column and eluted in the buffer containing 20 mM Tris-HCl, pH 7.4, and 150 mM NaCl. The purified complex was concentrated to 15 mg ml^−1^ for crystallization. Diffraction-quality crystals were grown at 16 °C by the sitting-drop vapour diffusion method using a 1:1 ratio of protein:reservoir solution. The reservoir solution contains 0.1 M sodium cacodylate (pH 6.5), 40% v/v MPD and 5% w/v PEG 8000. For data collection, crystals were transferred into reservoir solution before being cryo-preserved in liquid nitrogen. The diffraction data were collected at the National Facility for Protein Science in Shanghai (beamline BL18U1). Data were processed using autoPROC (*55*). The crystal structure was determined by molecular replacement using the program Phaser (*56*). The model of DDR2-DS domain in the published structure (PDB: 2WUH) and models for the variable region and the constant region of HL2-Fab generated by AlphaFold3 (*57*) were used as search models. The structure model was then manually adjusted in Coot (*58*) and refined using Phenix (*59*). The final structure was validated with the wwPDB server (*60*).

### Pull-down assay

To assess whether HL2 antibody binding affects the interaction between DDR2 and collagen, Protein A beads were first coated with the HL2 antibody via incubation in the binding buffer at 4°C for 30 min. Excess purified DDR2-DS protein was then added to the mixture, followed by incubation at 4°C for 30 min. After incubation, the beads were spun down and washed three times with the binding buffer to remove unbound DDR2. Next, type I collagen (Corning) was added, and the incubation was continued at 4°C for 30 min. Following a final wash step, protein complexes were eluted with 0.1 M Gly-HCl (pH 3.0). Eluted samples were analyzed by immunoblotting using an anti-His tag antibody (1:5,000; TransGen Biotech) and an anti-collagen antibody (1:1000; Selleck Chemicals).

To examine the interaction between HL2 antibody and DDR2-DS, an additional pull-down assay was performed as follows: 20 µg of the HL2 antibody was mixed with excess DDR2-DS or mutant protein in binding buffer and incubated on ice for 30 min. This mixture was then incubated with Protein A beads at 4°C for another 30 min with mild rotation. Subsequently, the beads were collected by centrifugation and washed three times with the binding buffer. Proteins retained on the beads were eluted using 0.1 M Gly-HCl (pH 3.0). Eluted samples were analyzed by non-reducing SDS-PAGE and Coomassie Brilliant Blue (CBB) staining.

### Antibody internalization assay

DDR2-overexpressing 293T cells were maintained in DMEM supplemented with 10% fetal bovine serum and 1% penicillin/streptomycin. For internalization assays, cells were seeded onto poly- L- lysine-coated coverslips (for immunofluorescence) or into 12-well plates (for flow cytometry) and allowed to adhere overnight. Cells were then incubated with HL2 antibody (mouse monoclonal, 0.8 μg/mL) at 37 °C in 5% CO₂. For immunofluorescence, treatment durations were 2, 4, 8, 12, and 24 h. For flow cytometric analysis, shorter time points of 0.5, 1, 2, 4, and 8 h were used to capture early internalization kinetics. Control cells were incubated in complete medium without HL2 antibody. After treatment, cells were washed twice with ice-cold PBS, fixed with 4% paraformaldehyde for 15 min at room temperature, and permeabilized with 0.1% Triton X-100 for 10 min. Non-specific binding was blocked with 5% bovine serum albumin (BSA) for 1 h. Internalized HL2 antibody was detected using Alexa Fluor 488-conjugated goat anti-mouse IgG (1:500, Invitrogen) diluted in 1% BSA/PBS for 1 h at room temperature. Nuclei were counterstained with DAPI (1 μg/mL). Coverslips were mounted with ProLong Diamond Antifade Mountant. Images were acquired on a confocal microscope (Zeiss LSM 900) using a 63× oil-immersion objective. For parallel quantification, treated cells were harvested by gentle trypsinization at the specified time points (0.5, 1, 2, 4, 8 h), washed twice with PBS, and fixed with 4% PFA for 15 min. After permeabilization with 0.1% saponin in PBS containing 1% BSA for 10 min, cells were stained with the same Alexa Fluor 488-conjugated secondary antibody for 30 min at 4 °C in the dark. Fluorescence was measured using a flow cytometer (ACEA Biosciences), and data from at least 10,000 events per sample were analyzed with FlowJo software. Median fluorescence intensity (MFI) of the FITC channel was used to compare HL2 internalization across time points.

### Detection of HL2 in blood and brain homogenates

Blood was collected via retro-orbital puncture from deeply anesthetized mice and transferred into EDTA-coated tubes. The samples were centrifuged at 3000 × g for 10 minutes at room temperature. Subsequently, 20 µL of the clarified plasma supernatant was aliquoted for analysis. For brain tissue analysis, a 50 mg section of brain tissue was dissected and homogenized in 250 µL of ice-cold phosphate-buffered saline (PBS) using a mechanical tissue homogenizer. Homogenization was performed at 60 Hz for 5 minutes at −20°C. The resulting homogenate was centrifuged at 12000 × g for 15 minutes at 4°C to remove cellular debris, and 200 µL of the supernatant was collected for analysis. For the binding assay, 1 × 10⁶ 293T-DDR2 cells were incubated with the prepared plasma or brain homogenate samples on ice for 1 hour, and then the HL2 was detected by incubating the cells with an Alexa Fluor 488-conjugated goat anti-mouse IgG secondary antibody on ice for 40 minutes in the dark. Flow cytometric analysis was performed using a NovoCyte instrument.

### Statistical analysis

Statistical analyses were performed using GraphPad Prism 8 software. All results were presented as mean ± SEM. In the case of normally distributed data, one-way analysis of variance (ANOVAs) and least significant difference (LSD)-*t*-tests were used to compare groups of more than three. Paired *t*-tests were used to compare two samples. Spearman’s correlation coefficients were calculated for the correlation analysis. Significance threshold: *p* < 0.05 (two-tailed).

### Data availability

The crystal structure of the HL2-Fab in complex with DDR2-DS has been deposited in PDB with the accession code 23KC.

## Data Availability

All data produced in the present study are available upon reasonable request to the authors

## Acknowledgements

We thank Dr. Kongzhen Hu, the Department of Nuclear Medicine, Nanfang Hospital, Southern Medical University, for the provision of CBP-FITC and CBP-NOTA. We gratefully acknowledge Feichuang Biomedical Technology (Guangzhou) Co., Ltd for their indispensable contribution to the screening of sdAb and the design of DDR2 *in vivo* imaging, which were pivotal to the success of this study. We thank the staff members at BL18U1 beamlines (https://cstr.cn/31129.02.NFPS.BL18U1) of the National Facility for Protein Science in Shanghai (https://cstr.cn/31129.02.NFPS), for technical support in X-ray diffraction data collection and analysis. We also thank the Drug Discovery Platform of Guangzhou National Lab for help with crystal screening.

## Author contribution

J.S. and P.Y. designed and conceived the experiments. P.Y., X.C., L.M., Z.L., T.G., J.J., X.H., G.L., B.S., and C.G. conducted the experiments. P.Y., Jia.L., Y.P. and Ya.L. analyzed the data. X.C., P. R. and P.Y. contributed to the PET/CT imaging. Y.X., Jun.L. and Yun L. purified 1A12-mCherry and HL2. J.D. performed the immunofluorescence staining for human samples. Y.P. carried out the crystallography studies and pull-down assays. Ya.L. conceived and supervised the structural investigation, and drafted the structural analysis section of the manuscript with input from Y.P. J.S., P. Y., Jia.L., and Ya.L. wrote the paper.

## References and Notes

1. J. Hardy, Alzheimer’s Disease: Treatment Challenges for the Future. J Neurochem 169, e70176 (2025).

2. G. Fatima et al., Vascular and glymphatic dysfunction as drivers of cognitive impairment in Alzheimer’s disease: Insights from computational approaches. Neurobiol Dis 208, 106877 (2025).

3. N. Korte, R. Nortley, D. Attwell, Cerebral blood flow decrease as an early pathological mechanism in Alzheimer’s disease. Acta Neuropathol 140, 793–810 (2020).

4. N. Korte et al., Inhibiting Ca(2+) channels in Alzheimer’s disease model mice relaxes pericytes, improves cerebral blood flow and reduces immune cell stalling and hypoxia. Nat Neurosci 27, 2086–2100 (2024).

5. D. M. Lopes et al., Glymphatic inhibition exacerbates tau propagation in an Alzheimer’s disease model. Alzheimers Res Ther 16, 71 (2024).

6. M. H. Murdock et al., Multisensory gamma stimulation promotes glymphatic clearance of amyloid. Nature 627, 149–156 (2024).

7. A. MohanaSundaram, M. Mofatteh, G. M. Ashraf, D. Praticò, Glymphotherapeutics for Alzheimer’s disease: Time to move the needle. Ageing Res Rev 101, 102478 (2024).

8. H. Gong, H. Zhong, Y. H. Ma, X. L. Li, D. K. Zhang, Unraveling the role of discoidin domain receptors as an anti-fibrotic target in various organs: A comprehensive review. Int J Biol Macromol 322, 146739 (2025).

9. C. Zeltz, M. Kusche-Gullberg, R. Heljasvaara, D. Gullberg, Novel roles for cooperating collagen receptor families in fibrotic niches. Curr Opin Cell Biol 85, 102273 (2023).

10. P. Trono, F. Ottavi, L. Rosano, Novel insights into the role of Discoidin domain receptor 2 (DDR2) in cancer progression: a new avenue of therapeutic intervention. Matrix Biol 125, 31–39 (2024).

11. S. Ling et al., Discoidin domain receptor 2 signaling through PIK3C2α in fibroblasts promotes lung fibrosis. J Pathol 262, 505–516 (2024).

12. H. Zhao et al., Targeting of Discoidin Domain Receptor 2 (DDR2) Prevents Myofibroblast Activation and Neovessel Formation During Pulmonary Fibrosis. Mol Ther 24, 1734–1744 (2016).

13. F. J. Garcia et al., Single-cell dissection of the human brain vasculature. Nature 603, 893–899 (2022).

14. M. Hebron et al., Discoidin domain receptor inhibition reduces neuropathology and attenuates inflammation in neurodegeneration models. J Neuroimmunol 311, 1–9 (2017).

15. H. Mathys et al., Single-cell multiregion dissection of Alzheimer’s disease. Nature 632, 858–868 (2024).

16. E. C. B. Johnson et al., Cerebrospinal fluid proteomics define the natural history of autosomal dominant Alzheimer’s disease. Nat Med 29, 1979–1988 (2023).

17. F. Yue et al., Synthetic amyloid-β oligomers drive early pathological progression of Alzheimer’s disease in nonhuman primates. iScience 24, 103207 (2021).

18. P. Désogère et al., Type I collagen-targeted PET probe for pulmonary fibrosis detection and staging in preclinical models. Sci Transl Med 9, (2017).

19. S. B. Montesi et al., Type I Collagen-targeted Positron Emission Tomography Imaging in Idiopathic Pulmonary Fibrosis: First-in-Human Studies. Am J Respir Crit Care Med 200, 258–261 (2019).

20. B. Leitinger, A. P. Kwan, The discoidin domain receptor DDR2 is a receptor for type X collagen. Matrix Biol 25, 355–364 (2006).

21. N. Singh, B. Das, J. Zhou, X. Hu, R. Yan, Targeted BACE-1 inhibition in microglia enhances amyloid clearance and improved cognitive performance. Sci Adv 8, eabo3610 (2022).

22. N. Y. Wang et al., Ferulic Acid Ameliorates Alzheimer’s Disease-like Pathology and Repairs Cognitive Decline by Preventing Capillary Hypofunction in APP/PS1 Mice. Neurotherapeutics 18, 1064–1080 (2021).

23. J. Ratelade et al., Reducing Hypermuscularization of the Transitional Segment Between Arterioles and Capillaries Protects Against Spontaneous Intracerebral Hemorrhage. Circulation 141, 2078–2094 (2020).

24. D. Liu et al., Regulation of blood-brain barrier integrity by Dmp1-expressing astrocytes through mitochondrial transfer. Sci Adv 10, eadk2913 (2024).

25. J. J. Iliff et al., A paravascular pathway facilitates CSF flow through the brain parenchyma and the clearance of interstitial solutes, including amyloid β. Sci Transl Med 4, 147ra111 (2012).

26. E. A. Yamamoto et al., The perivascular space is a conduit for cerebrospinal fluid flow in humans: A proof-of-principle report. Proc Natl Acad Sci U S A 121, e2407246121 (2024).

27. O. Ichikawa et al., Structural basis of the collagen-binding mode of discoidin domain receptor 2. Embo j 26, 4168–4176 (2007).

28. F. Carafoli et al., Crystallographic insight into collagen recognition by discoidin domain receptor 2. Structure 17, 1573–1581 (2009).

29. R. Shi et al., Tropism-shifted AAV-PHP.eB-mediated bFGF gene therapy promotes varied neurorestoration after ischemic stroke in mice. Neural Regen Res 21, 704–714 (2026).

30. Y. Chen et al., Targeting amyloid-β pathology by chimeric antigen receptor astrocyte (CAR-A) therapy. Science 391, eads3972 (2026).

31. S. Haas et al., Functional PET/MRI reveals active inhibition of neuronal activity during optogenetic activation of the nigrostriatal pathway. Sci Adv 10, eadn2776 (2024).

32. J. Osorio et al., Increased lung [(18)F]-FDG uptake in chronic thromboembolic pulmonary hypertension with distal involvement. Eur Respir J 66, (2025).

33. Y. Wang et al., Advance and Prospect of Positron Emission Tomography in Alzheimer’s disease research. Mol Psychiatry 30, 4899–4909 (2025).

34. G. B. Frisoni et al., New landscape of the diagnosis of Alzheimer’s disease. Lancet 406, 1389–1407 (2025).

35. T. Ren et al., Discoidin domain receptor 2 (DDR2) promotes breast cancer cell metastasis and the mechanism implicates epithelial-mesenchymal transition programme under hypoxia. J Pathol 234, 526–537 (2014).

36. S. C. Chen, B. W. Wang, D. L. Wang, K. G. Shyu, Hypoxia induces discoidin domain receptor-2 expression via the p38 pathway in vascular smooth muscle cells to increase their migration. Biochem Biophys Res Commun 374, 662–667 (2008).

37. H. M. Kim et al., Glycation-mediated tissue-level remodeling of brain meningeal membrane by aging. Aging Cell 22, e13805 (2023).

38. H. E. Jones et al., Meningeal origins and dynamics of perivascular fibroblast development on the mouse cerebral vasculature. Development 150, (2023).

39. L. Liu et al., Matrix-transmitted paratensile signaling enables myofibroblast-fibroblast cross talk in fibrosis expansion. Proc Natl Acad Sci U S A 117, 10832–10838 (2020).

40. S. C. Ugbaja, M. M. Lawal, H. M. Kumalo, An Overview of β-Amyloid Cleaving Enzyme 1 (BACE1) in Alzheimer’s Disease Therapy: Elucidating its Exosite-Binding Antibody and Allosteric Inhibitor. Curr Med Chem 29, 114–135 (2022).

41. A. V. Andjelkovic et al., Blood-Brain Barrier Dysfunction in Normal Aging and Neurodegeneration: Mechanisms, Impact, and Treatments. Stroke 54, 661–672 (2023).

42. S. Zhang et al., Experimental chondrocyte hypertrophy is promoted by the activation of discoidin domain receptor 2. Mol Med Rep 10, 1543–1548 (2014).

43. T. Butler et al., Choroid Plexus Calcification Correlates with Cortical Microglial Activation in Humans: A Multimodal PET, CT, MRI Study. AJNR Am J Neuroradiol 44, 776–782 (2023).

44. Z. Ruzi, K. Bozorov, L. Nie, J. Zhao, H. Akber Aisa, Discovery of novel (E)-1-methyl-9-(3-methylbenzylidene)-6,7,8,9-tetrahydropyrazolo[3,4-d]pyrido[1,2-a]pyrimidin-4(1H)-one as DDR2 kinase inhibitor: Synthesis, molecular docking, and anticancer properties. Bioorg Chem 135, 106506 (2023).

45. M. Carracedo et al., The tyrosine kinase inhibitor nilotinib targets the discoidin domain receptor DDR2 in calcific aortic valve stenosis. Br J Pharmacol 179, 4709–4721 (2022).

46. A. Binrayes, C. Ge, F. F. Mohamed, R. T. Franceschi, Role of Discoidin Domain Receptor 2 in Craniofacial Bone Regeneration. J Dent Res 100, 1359–1366 (2021).

47. M. E. Pizzo et al., Transferrin receptor-targeted anti-amyloid antibody enhances brain delivery and mitigates ARIA. Science 389, eads3204 (2025).

48. G. D. Rabinovici et al., Updated appropriate use criteria for amyloid and tau PET: A report from the Alzheimer’s Association and Society for Nuclear Medicine and Molecular Imaging Workgroup. Alzheimers Dement 21, e14338 (2025).

49. M. Bors, R. Tomic, D. M. Perlman, H. J. Kim, T. P. Whelan, Cognitive function in idiopathic pulmonary fibrosis. Chron Respir Dis 12, 365–372 (2015).

50. H. Annaka, T. Nomura, H. Moriyama, Cognitive Function in Patients With Mild Idiopathic Pulmonary Fibrosis: A Case-Control Pilot Study. Occup Ther Health Care 39, 397–411 (2025).

51. N. D’Ambrosi, S. Apolloni, Fibrotic Scar in Neurodegenerative Diseases. Front Immunol 11, 1394 (2020).

52. F. A. Wolf, P. Angerer, F. J. Theis, SCANPY: large-scale single-cell gene expression data analysis. Genome Biol 19, 15 (2018).

53. I. Korsunsky et al., Fast, sensitive and accurate integration of single-cell data with Harmony. Nat Methods 16, 1289–1296 (2019).

54. X. Qiu et al., Reversed graph embedding resolves complex single-cell trajectories. Nat Methods 14, 979–982 (2017).

55. C. Vonrhein et al., Data processing and analysis with the autoPROC toolbox. Acta Crystallogr D Biol Crystallogr 67, 293–302 (2011).

56. A. J. McCoy et al., Phaser crystallographic software. J Appl Crystallogr 40, 658–674 (2007).

57. J. Abramson et al., Accurate structure prediction of biomolecular interactions with AlphaFold 3. Nature 630, 493–500 (2024).

58. P. Emsley, K. Cowtan, Coot: model-building tools for molecular graphics. Acta Crystallogr D Biol Crystallogr 60, 2126–2132 (2004).

59. P. V. Afonine et al., Towards automated crystallographic structure refinement with phenix.refine. Acta Crystallogr D Biol Crystallogr 68, 352–367 (2012).

60. H. Berman, K. Henrick, H. Nakamura, Announcing the worldwide Protein Data Bank. Nat Struct Biol 10, 980 (2003).

